# The risk of drug resistance during long-acting antimicrobial therapy

**DOI:** 10.1101/2021.07.10.21260044

**Authors:** Anjalika Nande, Alison L. Hill

**Affiliations:** Program for Evolutionary Dynamics, Harvard University, Cambridge, MA 02138; Institute for Computational Medicine, Johns Hopkins University, Baltimore, MD 21218

## Abstract

The emergence of drug resistance during antimicrobial therapy is a major global health problem, especially for chronic infections like HIV, hepatitis B and C, and TB. Sub-optimal adherence to long-term treatment is an important contributor to resistance risk. New long-acting drugs are being developed for weekly, monthly, or less frequent dosing to improve adherence, but may lead to long-term exposure to intermediate drug levels. In this study we analyze the effect of dosing frequency on the risk of resistance evolving during time-varying drug levels. We find that long-acting therapies can increase, decrease, or have little effect on resistance, depending on the source (pre-existing or de novo) and degree of resistance, and rates of drug absorption and clearance. Long-acting therapies with rapid drug absorption, slow clearance, and strong WT inhibition tend to reduce resistance risks due to partially resistant strains in the early stages of treatment even if they don’t improve adherence. However, if subpopulations of microbes persist and can reactivate during suboptimal treatment, longer-acting therapies may substantially increase the resistance risk. Our results show that drug kinetics affect selection for resistance in a complicated manner, and that pathogen-specific models are needed to evaluate the benefits of new long-acting therapies.

## Introduction

In recent decades, highly effective drugs have helped reduce morbidity and mortality of chronic viral infections, like those caused by the human immunodeficiency virus (HIV) [1] and hepatitis B (HBV) [2] and C (HCV) [3] viruses. However, drug resistance can evolve rapidly within individual hosts due to the large population sizes and high replication and mutation rates of many viruses, rendering treatments ineffective [4]. Similar problems complicate treatment for chronic bacterial infections like tuberculosis (TB) [5]. The long treatment courses (months – years) required for chronic infections increase the opportunity for pathogens to adapt.

Effective treatments can also fail due to non-adherence (missed doses). Typical rates of adherence for long-term medications are between 50 − 75% [6], which may be far less than what is needed for effective treatments. For some HIV antiretroviral therapies, studies have estimated that patients need near perfect (> 95%) adherence for viral suppression [7], while for HBV, adherence levels > 80% have been associated with an approximately 90% reduction in the rate of treatment failure [8].

Long-acting drugs are being developed to help address the problem of imperfect adherence [9]. For example, a two-drug injectable treatment regimen (cabotegravir/rilpivirine) was recently approved for treating HIV, and with a multi-week half-life it is administered only once every 4 or 8 weeks as opposed to current daily dosing [10, 11]. Large investments are being made in developing long-acting treatments for HCV and TB, and prophylaxis for TB and malaria [12, 13], with some success already in animal models [14–16]. Long-acting lipoglycopeptide antibiotics are already available to treat bacterial skin infections [17]. Monoclonal antibodies are an emerging treatment for infectious diseases which can be engineered to have long half-lives [18]. While long-acting therapies are likely to increase overall patient adherence, their affect on resistance is unknown. Many studies have shown that sub-optimal adherence to daily pills contributes to resistance [19–24], but it is also possible that the long-term exposure to intermediate drug levels between doses of a long-acting drugs could facilitate the evolution of resistance [25, 26]. The goal of this study is to examine the role of drug dosing kinetics on the risk of resistance.

Drug resistance can arise from two sources: mutants that exist prior to treatment initiation or those that are produced during treatment [27, 28]. The relative contribution of these sources towards resistance is difficult to separate experimentally, but has been thoroughly investigated in a generalized model of intra-host viral dynamics for a completely resistant mutant in the presence of constant drug efficacy [29]. However, recent work on evolution in fluctuating environments [30] shows that the fate of mutants that are under time-dependent selection pressures can’t necessarily be predicted by the time-averaged selective effect alone; suggesting the effect of drug kinetics should be important. Numerous studies have integrated pharmacokinetics into mathematical models of infection dynamics [31–42], and have shown that the likelihood of generating and selecting for drug resistance depends on fluctuating drug levels. However, no studies have systematically studied the impact of changing a drug profile, as will occur with the reformulation of drugs into long-acting therapies.

In this study we expand previous models to analyze the effect of drug kinetics on the evolution of resistance, focusing specifically on the frequency of drug dosing. We incorporate competition between wild-type and resistant strains, the fitness costs and benefits of resistance, pharmacologically-relevant drug kinetics, and treatment adherence. We consider pre-existing and rescue (de novo) mutations as well as mutations arising from reactivation of latent infection. This framework allows us to determine the conditions under which long-acting therapy promotes versus inhibits the development of drug resistance.

## Model

To understand the impact of drug kinetics on the evolution of resistance, we used a stochastic model of viral dynamics within individual hosts [43] (see Supplementary Methods for equations). This model (**Figure 1A**) describes the interactions between target (uninfected) cells, drug-susceptible wild-type virus (WT), and a drug-resistant virus strain, in the presence of treatment. While this model was designed for chronic viral infections, our results are generalizable to other infections with density-dependent growth and direct-acting, infection-blocking therapeutics (e.g. [38, 39]). For now, we assume that the resistant mutant is generated from the WT by a single point mutation. We discuss the implications of more complex mutational pathways in the Supplement.

**Figure 1:**
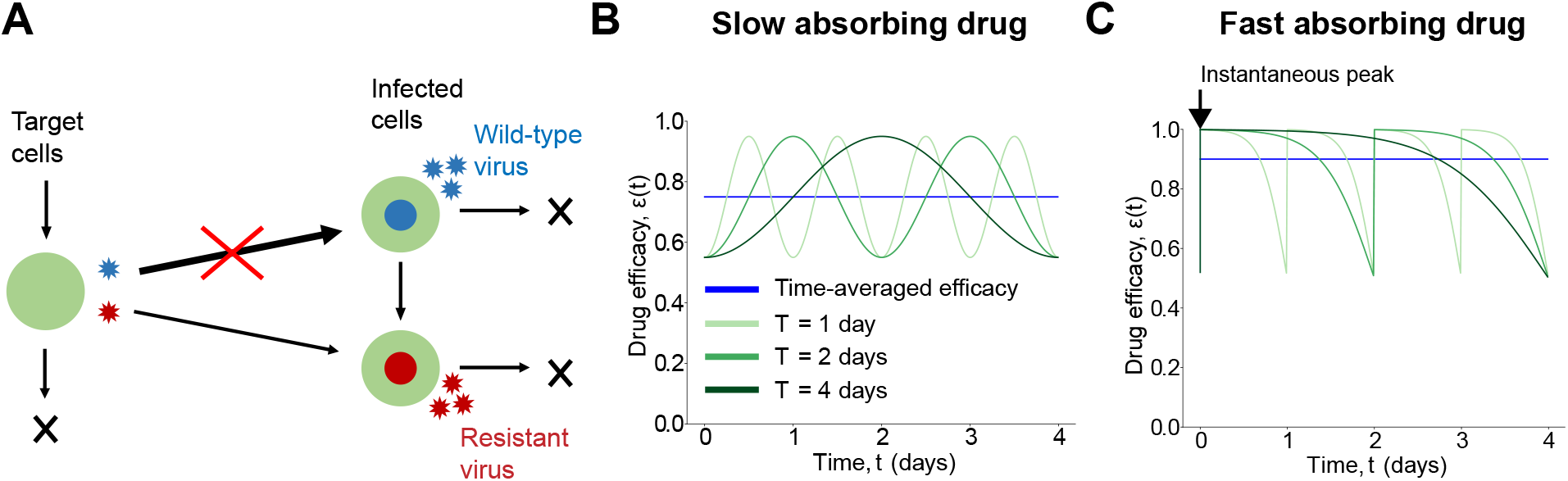
Model schematic and example time-dependent drug efficacy profiles. A) Schematic of the viral dynamics model consisting of uninfected target cells (green) and cells infected with either the wild-type (WT) (blue) or drug-resistant virus (red). We assume treatment blocks infection with the WT (red cross), while the resistant strain can still (at least partially) infect cells. The resistant virus is assumed to have a fitness cost, so in the absence of treatment, the WT is more infectious and dominates the population. The resistant strain can be produced via mutation from the WT. Time-dependent drug efficacy *ϵ*(*t*) under the B) slow and C) fast absorbing drug models. Green curves correspond to drug profiles with the same time-averaged (blue line), maximum and minimum efficacy, but with different dosing periods, *T*.

Like most infection models, pathogen fitness can be encapsulated by the basic reproductive ratio, *R*_0_, which is a composite of multiple individual parameters (Suppl. Methods). *R*_0_ is defined as the average number of new infected cells produced in a single replication cycle by one infected cell in an otherwise susceptible population [43]. A viral strain *i* can establish infection only if 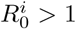. When there are multiple strains with 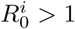, competitive exclusion occurs and only the strain with highest *R*_0_ can sustain high-level infection. We assume that resistance is accompanied by some fitness cost, 0 ≤ *s <* 1. Consequently, in the absence of treatment, 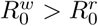, and infection is predominantly with the WT strain. The resistant strain is maintained in the population at low levels by a mutation-selection balance (**Figure S1**), which may lead to pre-existing resistance.

We assume that treatment reduces the infection rate by 1 − *ϵ*(*t*), where 0 *< ϵ* (*t*) *<* 1 is the time-varying drug efficacy. The drug is less efficacious against the resistant strain (*ϵ*_*r*_ *< ϵ*_*w*_). We are interested in a regime where an established WT infection is suppressed by treatment (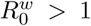 and 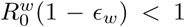), whereas the resistant strain is not 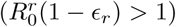. We consider two example drug efficacy models (Suppl. Methods): a ‘slow absorbing’ model in which drug absorption is gradual, such that the peak efficacy occurs in the middle of the dosing interval (**Figure 1B**), and a ‘fast absorbing’ model in which drug efficacy peaks immediately after dose administration and then decays continuously until the next dose (**Figure 1C**). This choice allows us to investigate the role played by the relative rates of drug absorption and clearance on the risk of resistance which is important for long-acting drugs as they are expected to have large differences in their absorption profiles [44]. With each of these models, we systematically varied the dosing interval while keeping the average efficacy the same (as well as the peak and trough efficacy). This is meant to mimic the scenario under which long-acting therapies are being developed: we assume that if the drug decay can be slowed down by some amount, the dosing interval is extended by the same amount.

Conceptually, there are two possible ways drug resistance can emerge to therapy [27, 29]. Drug resistant strains may pre-exist at the start of therapy, since they are continually produced by the WT strain and exist at mutation-selection balance. Once the WT infection is suppressed by therapy, the resistant strain has less competition for target cells (the “resource”), and can potentially reestablish infection. However, establishment is not guaranteed: the population of pre-existing resistant mutants can be very small and subject to stochastic extinction. Resistant infection can also be established by mutants produced by residual WT replication during therapy (referred to as “rescue mutants” following [29], a term borrowed from population genetics literature [45]), or later on in the treatment course due to reactivation of latent infected cells (for certain infections). These mutants are also subject to stochastic extinction. The establishment probability of a resistant infection is therefore a result of complicated, time-varying birth-death dynamics and multiple model parameters (e.g. rate of target cell production, cost of resistance, see Supplementary Information for further details). In the following sections we analyze the effects of drug dosing intervals on this probability for different sources of mutants.

## Results

### Resistant mutants existing prior to treatment

We first examined the impact of the dosing interval on the evolution of resistance from pre-existing mutants, which depends both on their number at the time of treatment initiation and the subsequent establishment probability (*p*_est_) of each of them. Since only the latter depends on the treatment course, we calculated the establishment probability for a single pre-existing resistant mutant in the presence of treatment, from traditional daily therapy to long-acting multi-monthly intervals with the same average, maximum, and minimum efficacy (Suppl. Methods). We first examined the case of a fully-resistant mutant (a strain completely unaffected by the drug level, purple curves in **Figures 2B,E**). For the slow absorbing drug profile, longer dosing intervals (i.e. long-acting therapy) always lead to lower probabilities of resistant mutants establishing. In contrast, for the fast absorbing drug profile, increasing the dosing period makes it more likely the resistant strain will establish. However in either case, the differences in *p*_est_ were minimal. We next looked at the case where the mutant is only partially resistant (green curves in **Figures 2B,E**). Overall the establishment probability was lower and more sensitive to the drug dosing frequency. When drug was absorbed more slowly between doses, longer dosing intervals only led to slightly lower establishment probability up to a certain maximal dose period (∼ monthly for the parameters we used), and then further increasing the period leads to even higher establishment probabilities than daily dosing. When drug was absorbed quickly, less frequent dosing decreased the establishment probability. Therefore, the effect of increasing dosing intervals on the risk of resistance due to pre-existing mutants seems to depend strongly on both the degree of resistance and on the details of the drug kinetics curve.

**Figure 2:**
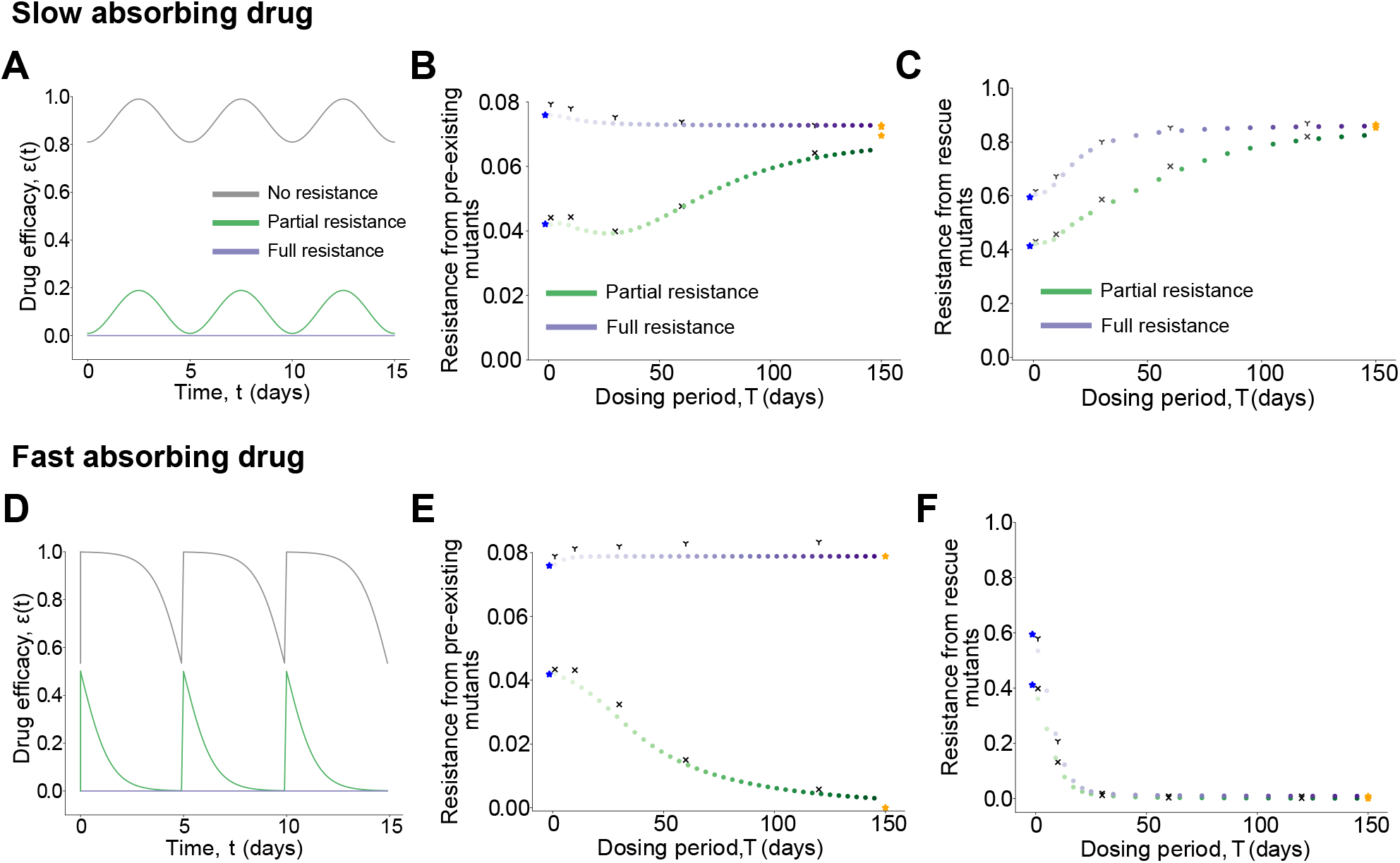
Effect of drug dosing intervals on the establishment of resistance due to pre-existing and rescue mutants. Results from numerical simulations for the slow (top row) and fast (bottom row) absorbing drug models. A) and D) Drug efficacy *ϵ*(*t*) versus time for the WT and resistant strain. B) and E) Establishment probability for one pre-existing resistant mutant as a function of the dosing interval. C) and F) Probability that at least one rescue mutant is produced as a function of the dosing interval. Each dot represents the establishment or rescue probability in the presence of an oscillatory efficacy with a fixed period. Colors go from light to dark with increasing dosing intervals keeping constant time-averaged efficacy. Results are for mutants unaffected (purple) and affected (green) by the drug. Stars correspond to the probabilities for constant efficacies: time-averaged efficacy (blue) and efficacy at *t* = 0 (orange). The overlaid black marks are results from stochastic simulations of the model (Suppl. Methods).

We identified an important timescale in the system that helps explain the complex relationship observed between the dosing frequency and the risk of pre-existing resistance establishing. When therapy begins, the resistant strain has a selective advantage over the WT, but the establishment probability is extremely low until the WT strain is sufficiently suppressed to remove competition for the limited resource they share (i.e., target cells). Each individual mutant has a finite lifespan, and the lineage of this strain must survive this time period to make establishment likely.

We call this timescale the establishment time frame (ETF), and we discuss its derivation and dependence on model parameters in the Supplement (**Figure S2**, Suppl. Methods). The effective drug levels during the ETF are the most relevant for resistance risk, explaining the observation of two limiting cases: when dosing is frequent, the drug undergoes many cycles during this time frame and the establishment probability is well-approximated by assuming constant efficacy at the time-averaged value (see blue stars in **Figures 2B,E**). When drug dosing is very infrequent (i.e. long-acting therapy), the establishment probability approaches the value predicted from the earliest drug levels (orange stars in **Figures 2B,E**).

Divergent results for partially vs fully-resistant mutants can also be explained by selection forces acting during the ETF. Higher drug efficacy during this time period indirectly promotes the resistant strain by suppressing WT competition, but if the mutant is not completely resistant, the drug also causes some partial direct dose-dependent inhibition. Thus, for a fully-resistant mutant, establishment is more likely for drug profiles with higher effective efficacy during the ETF (**Figures 2B,E**), whereas for a partially-resistant mutant the opposite is true when the inhibition by the drug is the dominant selection force (**Figures 2B,E**). The trends described here are robust to the choice of parameter values, and we discuss this in detail in the Supplementary Discussion along with **Figures S3,S4**.

To summarize, when partial inhibition of the resistant strain during therapy is the dominant selection force, long-acting drugs tend to increase the risk of resistance due to pre-existing mutants when the drug absorption is slow, and decrease the risk when drug absorption is fast. The converse is true when the degree of resistance is high and competition with the WT dominates instead.

### Resistant mutants produced during treatment

Next, we examined the effect of dosing frequency on the evolution of resistance due to *de novo* mutants produced during treatment. To contribute to treatment failure, a resistant lineage must first be produced by mutation during residual WT replication despite therapy and then have a strong enough selective advantage to escape stochastic extinction. We computed the probability that at least one such “rescue mutant” is produced during treatment (Suppl. Methods), and analyzed how it was affected by varying the frequency of drug dosing while keeping the average, maximum, and minimum drug efficacy fixed (**Figures 2C,F**).

We found that for slow drug absorption, long-acting drugs tend to increase the risk of resistance (**Figures 2C**). In contrast, for fast drug absorption, longer acting drugs reduce the risk of resistance (**Figures 2F**). These trends are independent of the degree of resistance of the mutant strain, and are robust to variations in other parameters (see Suppl. Discussion and **Figures S5,S6** for details).

The establishment time frame also plays an important role in explaining the risks of de novo resistance emerging, but through different mechanisms. The rate at which rescue mutants are produced depends on the product of the rate of mutant production from residual WT replication before infection is controlled, which is increased for lower drug efficacy during the ETF, and the establishment probability of these newly-generated mutants, which may be increased for either higher or lower drug efficacy depending upon the degree of resistance of the mutant strain (as was seen in the case of pre-existence).

Our findings suggest that the impact of drug kinetics on the rate of mutant production dominates the effect on mutant selection (and establishment probability). Drug profiles with higher efficacy shortly after treatment begins (e.g. frequently dosed slow-absorbing drugs or long-acting fast-absorbing drugs) are associated with lower risks of resistance from rescue mutants. There are two limiting cases again: when dosing is frequent, the rescue probability depends upon the time-averaged drug efficacy, whereas in the limit of very infrequent dosing it depends upon the initial drug efficacy.

In summary, long-acting therapy increases the risk of resistance due to rescue mutations when drug absorption is slow, and decreases it when drug absorption is fast.

### Resistant mutants produced due to latency reactivation

The infection model considered so far is applicable to infections like HCV that do not have persistent infected cell populations [46]. As a result, the infection tends to be cleared quickly as long as resistance doesn’t emerge, which is why the early drug kinetics play an out-sized role in the risk of resistance. Persistence, however, is a characteristic of many chronic infections (e.g. HIV, HBV, TB, some types of malaria) [5, 47] that makes it difficult to design effective treatment strategies to completely clear the infection. It is also an important source of resistance. Reactivation events from this pool (“latent reservoir”) of persistent infected cells can lead to the introduction of resistant mutants that establish infection [35, 48, 49]. The rate at which resistant mutants are produced and go on to establish infection (“rescue” due to reactivation) is dependent upon the details of the drug kinetics and hence, we expect it to be influenced by the drug dosing frequency. With this in mind, we investigated how long-acting therapy affects the emergence of a resistant infection due to the reactivation of latent infected cells.

We modeled two ways in which resistant mutants can be introduced during a persistent infection (Suppl. Methods) - reactivation of a latently-infected lineage that is already resistant (we assume that a fraction of the latent population given by mutation-selection balance was resistant at the time it was seeded), or reactivation and subsequent replication of WT infection that generates a resistant mutant (**Figure 3A**). Summing up these possibilities, we calculated the average probability of rescue per latency reactivation event (Suppl. Methods).

**Figure 3:**
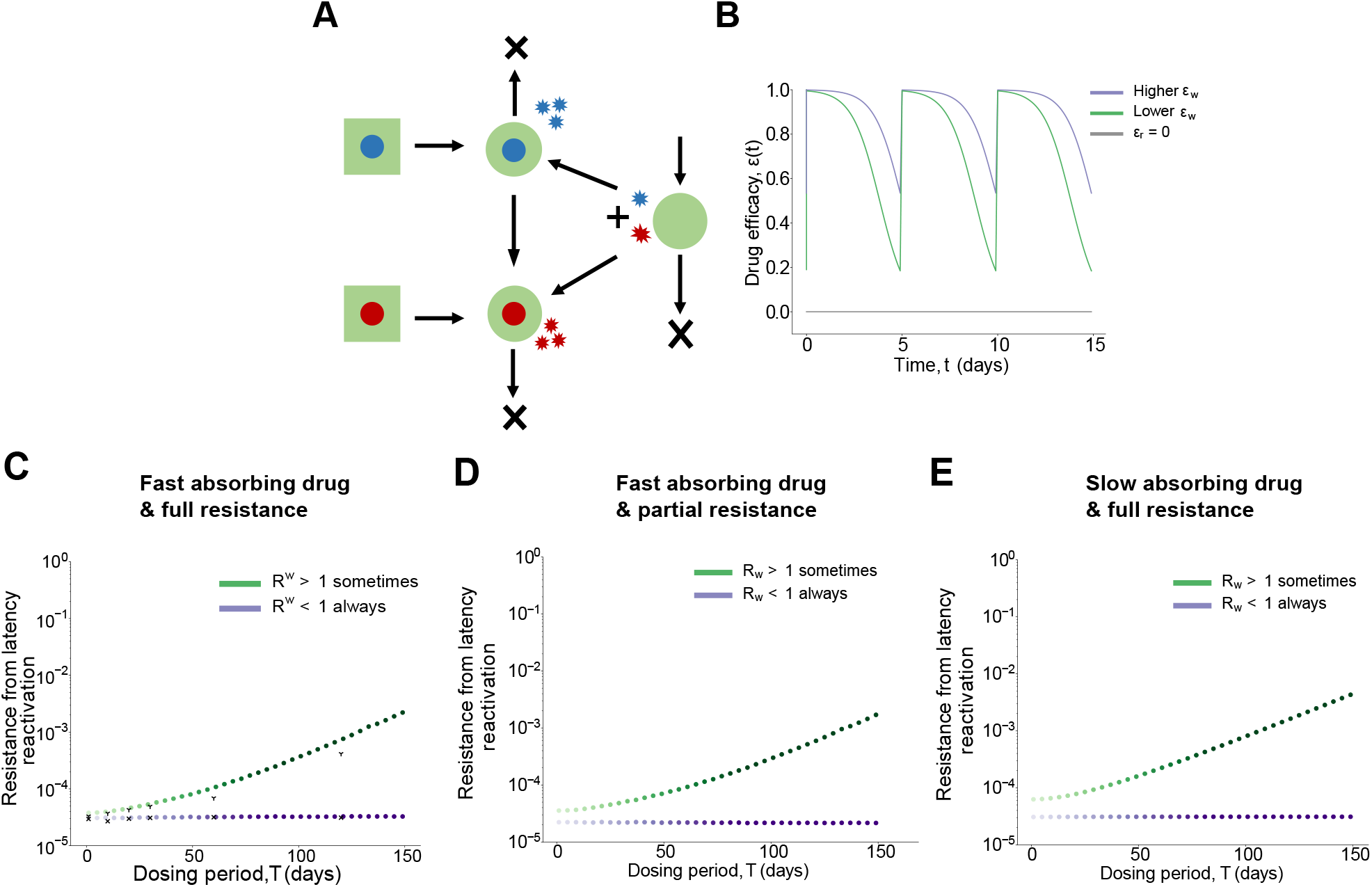
Effect of drug dosing intervals on the establishment of resistance due to latency reactivation. A) Model schematic showing the two ways in which resistant mutants can be produced via a latent infection (square). B) Example drug efficacy *ϵ*(*t*) versus time curves for WT (*ϵ*_*w*_) and resistant (*ϵ*_*r*_) strains under the fast absorbing drug model. The purple curve corresponds to a high drug efficacy with *R*^*w*^ *<* 1 for all points of the drug cycle. *R*^*w*^ > 1 for ∼23% of each period for the lower efficacy drug (green). C-E) Average probability of rescue per latency reactivation event as a function of the drug dosing interval for different drug efficacy profiles and degrees of resistance. Colors darken with increasing T keeping constant time-averaged efficacy. Black marks in C) denote results of the stochastic simulation of the model. See Suppl. Methods for details on the parameter values.

We found that depending upon the degree of WT inhibition between doses, long-acting therapies are associated with the same or increased risk of resistance as compared to frequent dosing (**Figure 3C**). When the drug efficacy is high enough that the WT is suppressed (*R*^*w*^ *<* 1) for the entire the drug cycle, the average rate of rescue has no dependence upon the dosing interval (purple curve in **Figure 3C**). In the absence of WT replication, resistance can arise only via mutants that pre-exist in the latent reservoir. Since we assume the rate of reactivation is constant and independent of the drug kinetics, the risk of resistance in this case does not depend upon the drug dosing frequency. However, when the drug efficacy is lower and the WT can replicate during the times of lowest drug levels (*R*^*w*^ > 1), despite still being suppressed overall (⟨*R*^*w*^⟩ *<* 1), the rate of rescue has a strong dependence upon the dosing intervals and increases for longer acting drugs (green curve in **Figure 3C**). The slow decay of longer acting drugs leads to more time in the non-suppressive part of the dosing period, enabling the WT to undergo multiple rounds of replication before eventually getting suppressed by the next dose, and increases the chance that a resistant mutant is produced and establishes infection. These results are irrespective of the degree of resistance of the mutant strain (**Figure 3D**) and the drug absorption rates (**Figure 3E**).

In summary, during a persistent infection, long-acting therapies are associated with an increased risk of resistance if there is sub-optimal WT suppression in the lowest parts of the drug cycle.

### Impact of non-adherence on resistance for different dosing intervals

The previous sections show how drug kinetics influence the risk of resistance when there is perfect adherence to treatment. In reality, treatment adherence is known to be sub-optimal, especially for chronic infections, and is associated with the risk of resistance for daily therapies. We therefore evaluated how imperfect adherence to therapy influences the relationship between drug dosing intervals and the risk of resistance. We assumed each scheduled dose was taken or missed randomly and independently with a probability given by the adherence level (Suppl. Methods) and analyzed the effect of dosing intervals on the probability of a resistant infection due to the different sources of mutations, for varying levels of adherence (**Figure 4**). While in reality low enough adherence levels can allow for treatment failure to occur *without* resistance, just due to rebound of the WT strain, we were not interested in this effect and so ensured that the system stayed in the regime where the WT has a lower fitness than the resistant strain and is suppressed by treatment.

**Figure 4:**
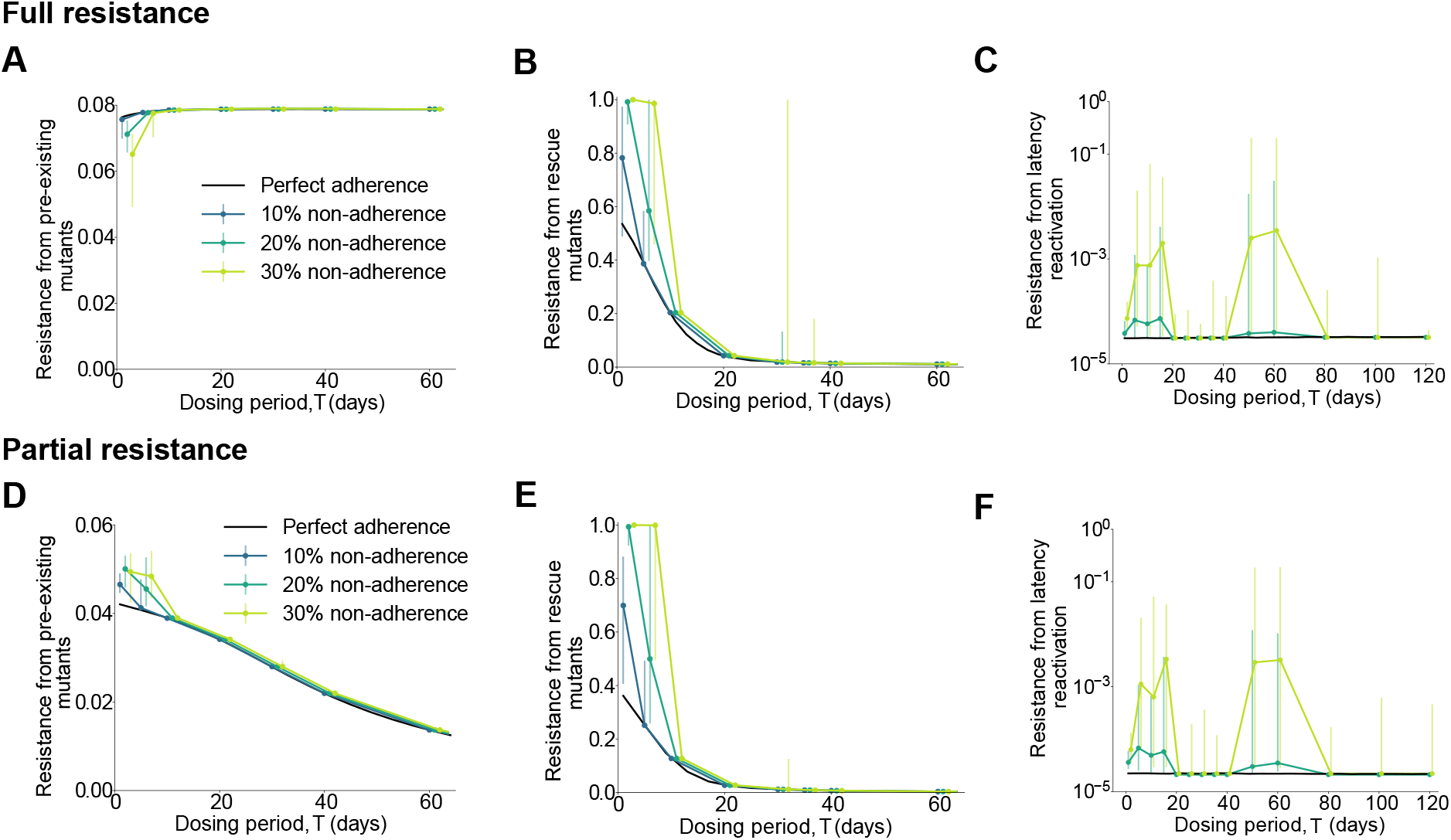
Effect of non-adherence on the establishment of resistance for different drug dosing intervals. Results for a mutant fully (top row) and partially (bottom row) resistant to treatment. A) and D) Establishment probability of pre-existing mutants. B) and E) Probability that at least one rescue mutant is produced. C) and F) Average probability of rescue per latency reactivation event. Results are for the fast absorbing drug model with the solid black line in each subplot corresponding to results for perfect adherence. Imperfect adherence results are medians of 100 iterations for pre-existence and rescue, and 500 iterations for latency. The error bars correspond to the interquartile range. X-axis positions are offset for ease of visualization.

We found that for both pre-existing and rescue mutants, long-acting drugs were more robust to the effects of treatment non-adherence, with the same observed overall trends relating the risk of resistance to the frequency of dosing as for perfect adherence (**Figures 4A,B,D and E**). Long-acting drugs are more robust in this case because the risk of resistance is mainly dependent upon the treatment strength during the establishment time-frame. The number of doses taken during this time-frame decreases as the dosing interval increases, reducing the chance one of them is missed. As by definition the first dose is always taken, the risk of resistance is the same as that for perfect adherence when the dosing interval is equal to or larger than the ETF. These results are independent of the specific model chosen for adherence (see Suppl. Methods and **Figure S7** for results with an alternative adherence model in which missed doses can be taken on any subsequent day, instead of waiting for the next scheduled dose).

In agreement with previous modeling work [35,39], we found that non-adherence generally - but not always - promotes the emergence of resistance. In the case of fully-resistant pre-existing mutants, lower adherence actually lowers the establishment probability (**Figure 4A**). With lower adherence, the WT infection is less effectively suppressed, which increases competition for target cells and makes it more difficult for the resistant strain to establish. This highlights the more general observation from other modeling studies [27, 38, 50] and experiments [51, 52] that an “aggressive” treatment (e.g., high initial drug levels) doesn’t necessarily lead to reduced resistance risk.

Depending upon the drug dosing frequency, imperfect adherence can significantly increase (by orders of magnitude) the risk of resistance during a persistent infection irrespective of the degree of resistance of the mutant strain (**Figures 4C,F**). The non-monotonic pattern of resistance risk vs dosing period and large error bars imply that the resistance risks are very sensitive to the state of the system at the time the mutant arises. When drugs are very long-acting (drug dosing period larger than ∼ 2 months), imperfect adherence doesn’t play much of a role – missing a dose causes WT infection to rebound without selecting for resistance. However, for shorter dosing periods the risk of resistance is predominantly increased, apart from an intermediate regime. This trend is a combined effect of the rate at which resistant mutants are produced from the WT and competition between the two strains for uninfected cells once the resistant mutants have been produced. The risk of resistance is highest in the regime where missing a dose leads to a burst of WT replication and mutant production, but where there is enough WT suppression overall that a resistant strain can establish infection. This is also why the overall rates of resistance are higher if we instead consider an alternative adherence model where individuals are given the chance to take a missed dose on any subsequent day, instead of waiting for the next scheduled dose (Suppl. Methods, Suppl. Discussion, **Figure S7**). Although on average the number of missed doses are equivalent in both models, the distribution of the number of days between doses is narrower in the second model and results in drug levels reaching highest-risk intermediate levels more frequently.

The results presented in this work correspond to a model where we assume that the resistant strain is one mutational step away from the WT. However, we find that the results also qualitatively hold for a more complex mutational pathway (see Suppl. Methods and **Figure S8** for more details).

To summarize, longer acting drugs are more robust to the effects of imperfect adherence when drug resistance is expected to arise due to mutants pre-existing or generated early on after the start of treatment. During a persistent infection, non-adherence to long-acting drugs can dramatically increase the risk of resistance occurring if latent infection reactivates compared to daily dosing.

## Discussion

New long-acting antimicrobial therapies are being developed to improve patient adherence and reduce treatment failure [9]. Although these drug formulations are likely to reduce missed doses, it is unclear whether this alone will lead to better treatment outcomes. Therapies with extended half-lives can lead to long-term exposure to intermediate drug levels, which could promote the evolution of drug resistance [25, 26]. A systematic study of the relationship between drug dosing kinetics and the risk of resistance is therefore essential to the development and optimization of new long-acting therapies.

In this study we found that the interplay between time-varying drug concentrations in the body and competing pathogen strains results in a complicated effect of drug dosing intervals on the evolution of resistance (**Table 1**). Using models of infection dynamics within hosts, we show that when resistance mutations pre-exist before treatment and confer complete resistance, long-acting therapies increase the resistance risk if drug absorption is fast and reduce it if drug absorption is slow. These trends can be reversed if pre-existing mutants confer only partial resistance. These opposing results suggest that in cases where complex resistance pathways exist, full disease and treatment-specific models that attempt to account for realistic fitness landscapes of resistance – such as the model developed by Raja et al. [53] or the one developed by Kirtane et al. [54] – will be needed to evaluate the risks of resistance and guide treatments. When the main source of resistance is mutants produced de novo via residual wild-type infection after treatment begins (“rescue”), then irrespective of the degree of resistance, long-acting drugs reduce the resistance risk if drug absorption is fast and increase it if drug absorption is slow. Our results demonstrate that resistance risk due to both pre-existing and rescue mutations is dominated by drug kinetics (treatment strength) and infection dynamics in a short initial time window after treatment begins, which we call the “establishment time frame”. It is worth noting that our analysis essentially amounts to calculating the fixation probability of a mutant under time-varying selection pressures, a complicated problem that has been studied in the field of population genetics. We relate our findings to previous work in the Supplement.

**Table 1:**
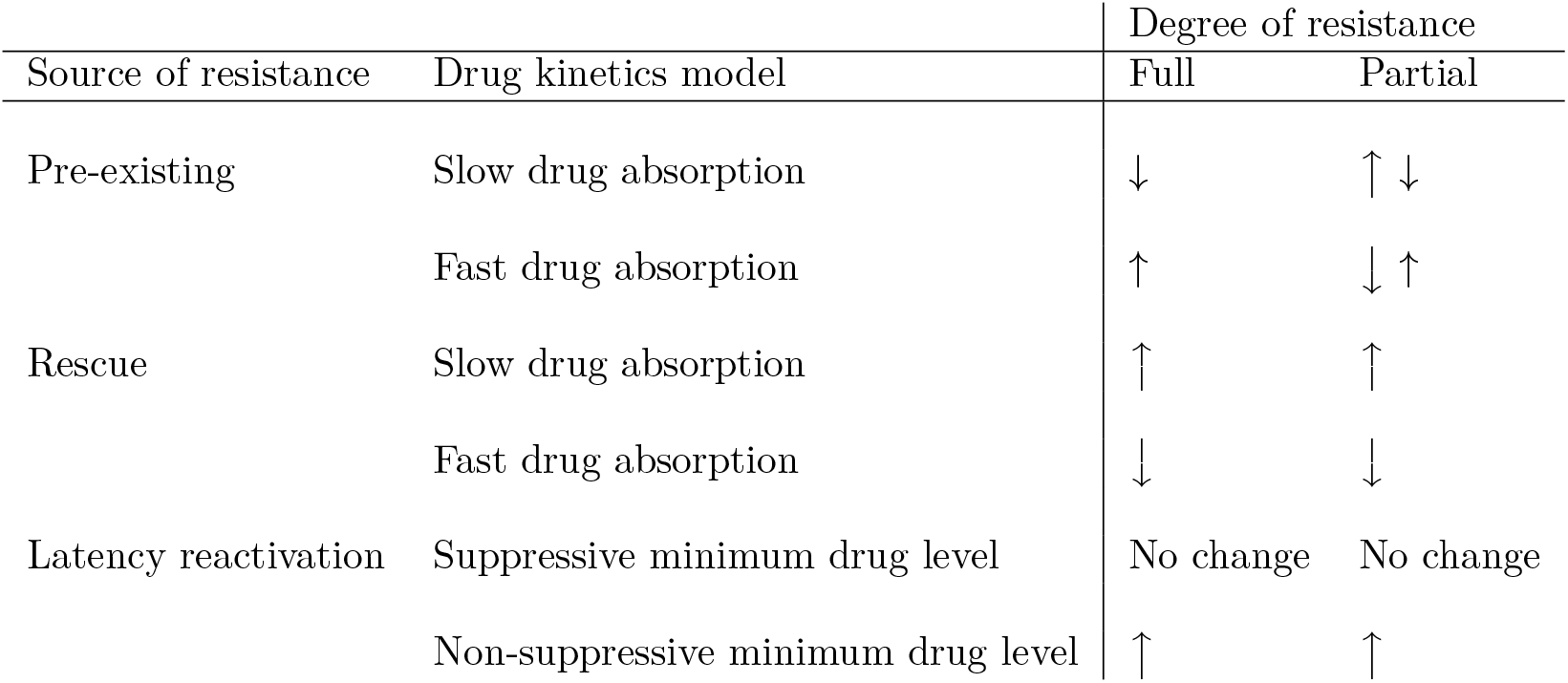
Summary of effect of long-acting drugs on the risk of resistance during therapy. “↑” means that the probability of drug resistance emerging during long-acting therapy is higher compared to daily dosing (with the same average, peak, and minimum levels), and “↓” means the opposite. Smaller sized arrows indicate minimal effect. For resistance arising from mutations that are either “pre-existing” at the time treatment starts or from “rescue” (de novo) mutations produced by residual replication before wild-type infection is suppressed, the speed of drug absorption (relative to clearance) is the important pharmacokinetic factor determining resistance risk. For resistance arising from reactivation of persistent infection long after therapy initiation, the inhibitory effect of the minimum drug level is the most important pharmacokinetic factor determining resistance risk (with a more minor role played by the drug absorption rate and degree of resistance). Results come from simulations with a specific infection model and parameter values, and while our analyses suggest trends are relatively general, the magnitude of the effect will change with model details (See Suppl. Discussion for more details).

| Source of resistance | Drug kinetics model | Degree of resistance |  |
| --- | --- | --- | --- |
|  |  | Full | Partial |
| Pre-existing | Slow drug absorption | ↓ | ↑ ↓ |
|  | Fast drug absorption | ↑ | ↓ ↑ |
| Rescue | Slow drug absorption | ↑ | ↑ |
|  | Fast drug absorption | ↓ | ↓ |
| Latency reactivation | Suppressive minimum drug level | No change | No change |
|  | Non-suppressive minimum drug level | ↑ | ↑ |

Treatment for infections such as HIV and TB can be compromised by persistent sources of infection which aren’t rapidly cleared by therapy. For resistance arising from reactivation of persistent reservoirs, early drug kinetics (and thus the absorption speed of long-acting therapies and the degree of resistance) become less important. Resistant mutants can arise at any time from pre-existing mutant populations in the persistor pool, or get generated during breakthrough replication of WT cells from the persistent population when drug levels are low. We found that during persistent infection, longer-acting therapies increase the risk of resistance if the drug isn’t fully suppressive during the lowest drug levels. Long-term exposure to low drug levels can allow WT populations that reactivate from latency to temporarily grow, significantly increasing the chance that resistant mutants are generated and expand. Individual physiological differences in drug absorption, distribution, metabolism, and excretion mean that the suppressive action of therapy can vary significantly across a population, and previous studies have stressed the need to individualize treatment regimens [32, 55]. Our findings suggest that this need might be even more acute for longer-acting therapies.

Imperfect treatment adherence is a major challenge for long-term antimicrobial therapy and can facilitate the emergence of resistance. While the development of long-acting therapies was motivated by the need to improve adherence, their robustness to missed doses has not been evaluated. Mathematical models have helped provide valuable insights into understanding the complex and at times surprising effects of treatment non-adherence [31–36, 39, 56]. Extending this work to long-acting therapies, we find that the effects of imperfect adherence depend on whether there is a persistent source of infection. In its absence, long-acting drugs are more robust to the effects of imperfect adherence compared to more frequent dosing, since it is less likely doses will be missed in the critical early time frame during which wild-type infection is cleared before resistant strains can be produced or effectively compete for resources. However, if there is a persistent population from which infection can reactivate during later periods of non-adherence, then some intermediate dosing frequencies lead to dramatically higher resistance risks.

Our model assumes a single well-mixed pathogen population. In reality, pathogen subpopulations can reside in tissues and organs where drug absorption and pathogen dynamics may be significantly altered [57–59]. Previous work has considered the consequences of spatial heterogeneity on treatment efficacy and the role it can play in facilitating the emergence of drug resistance [60–66]. In our analysis, the rate of drug absorption played an important role in determining the risk of resistance to long-acting therapies. Since drug absorption and clearance can differ between tissues, we suspect that the relationship between the frequency of dosing and the emergence of resistance could be more complex for compartmentalized infections.

The injectable combination cabotegravir/rilpivirine is the only long-acting antiretroviral therapy for HIV in late-stage of clinical trials, and after showing non-inferiority to the current standard daily oral dosing it was recently approved for use in many countries [10,11,67,68]. Our analysis provides some insight into what to expect long-term. In these trials, participants initiating therapy were given standard daily doses until viral suppression was achieved, after which they were randomly assigned to a longer dosing schedule (4 or 8 weeks) or kept at daily dosing as a control. Our analysis suggests that by switching to longer-acting therapy only after viral suppression is achieved, the risk of resistance from pre-existing or rescue mutations during the critical establishment time frame may be negated. HIV infection will still persist indefinitely in the the latent reservoir and may reactivate during periods of non-adherence, but results suggest that long-acting therapy will not be associated with an increased risk of resistance if drug levels are kept high enough in all individuals to ensure WT suppression even at the lowest points of the drug-cycle, which existing pharmacokinetic data supports [69, 70]. The rates of non-adherence for these monthly or bi-monthly injections has yet to be determined.

Long-acting drugs are also being considered for prophylactic use to improve adherence and increase effectiveness in preventing infection, for example for HIV and malaria [14,71,72]. For long-acting preventative therapy, there are concerns about the emergence of resistance due to long-term exposure of intermediate drug levels if prophylaxis is insufficient to prevent infection, or is initiated during an undiagnosed acute infection [73, 74]. Penrose et al [74] report an instance of infection with wild-type HIV and subsequent selection of a resistant virus due to persistent exposure to long-acting pre-exposure prophylaxis (PrEP). In addition, Radzio-Basu et al [73] show in a macaque model that initiating long-acting PrEP during acute simian-human immunodeficiency virus (SHIV) infection can frequently select mutations conferring resistance to therapy and are maintained for several months. Although we have not considered prophylactic use directly, some of our results seem qualitatively applicable. The risk of resistance if prophylaxis is unknowingly started during acute infection may be similar to our findings for the risk due to rescue mutations after therapy initiation. The risk of resistance during breakthrough infections is likely to depend on drug-kinetics in a similar way to that of reactivating persistent infection, but the chance of breakthrough infection itself may require a more specific model of transmission.

Our findings underscore the importance of explicitly modeling drug kinetics, since time-averaged drug efficacy is a poor proxy for resistance risk. Future work should focus on developing application-specific models that use data-driven pharmacodynamic, pharmacokinetic, and evolutionary parameters. Such models can help guide future dose-optimization and implementation tasks for long-acting therapies for globally important infectious diseases.

## Data Availability

Our code is open access and shared on Github

https://github.com/anjalika-nande/drugkinetics-resistance

## Acknowledgements

This work was supported by the Bill & Melinda Gates Foundation (OPP1148627). The computations in this paper were run on the FASRC Cannon cluster supported by the FAS Division of Science Research Computing Group at Harvard University. We thank Giovanni Traverso, Ameya Kirtane, Morgan Craig, Martin Nowak, Katharine Best, Madison Ski Krieger, Michael Desai, and John Wakeley for helpful feedback on this project.

## Data accessibility

Our code is open access on GitHub https://github.com/anjalika-nande/drugkinetics-resistance.

## Supplementary Information

### Supplementary Methods Infection dynamics model Before treatment

We start from a standard ordinary differential equation model of intra-host viral dynamics [29,43] consisting of uninfected target cells *x*, cells infected by the drug-sensitive wild-type (WT) strain *y*_*w*_, cells infected by the drug-resistant strain *y*_*r*_, free WT virions *v*_*w*_ and free resistant virions *v*_*r*_,

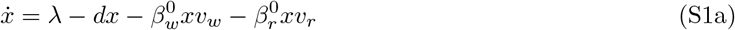

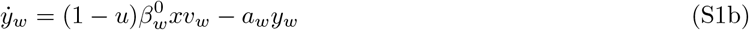

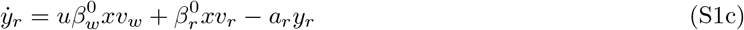

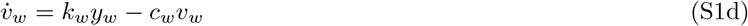

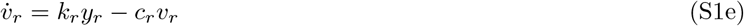

where *λ* is the rate of target cell production (assumed to be constant for simplicity); *d* is the natural death rate of target cells; 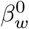 and 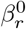 are the virus specific infectivities with the superscript indicating the value of the parameters in the absence of treatment; *a*_*w*_ and *a*_*r*_ are the death rates of the infected cells; *k*_*w*_ and *k*_*r*_ are the virion production rates; *c*_*w*_ and *c*_*r*_ are the free virion clearance rates; and *u* is the mutation rate.

Mutation is modeled according to the ‘stamping machine’ mode of replication [77, 78] and back mutation is ignored. The results discussed in this paper don’t depend upon when mutation occurs in a viral life-cycle and so we assume it occurs at the infection step without any loss of generality. Since we are modeling a system with only two strains, *u* represents the effective mutation rate. It encapsulates the effect of all genetic changes that lead to resistance. This simplification ignores the effect of multi-step mutational pathways. We briefly discuss the effects of higher point mutants in later sections. However, this simple two-strain model is known to be good approximation when the fitness of the intermediate strains is low [29].

The fitness of the strains is summarized by their basic reproduction ratio, *R*_0_. *R*_0_ is defined as the average number of new infected cells produced in a single replication cycle by an infected cell introduced into a population of uninfected cells [43]. The WT reproduction ratio in the absence of drugs is given by,

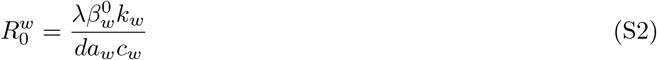

The resistant strain has some associated cost of resistance, *s*, that is incorporated such that 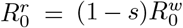. We simplify the above model further by using the common assumption that the dynamics of the free virions are much faster than those of the infected cells [79]. This leads to quasi-equilibrium relationships 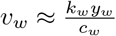 and 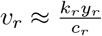 and the system **Equation S1** becomes,

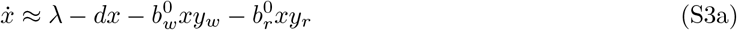

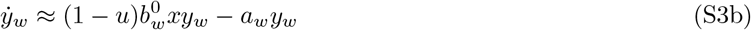

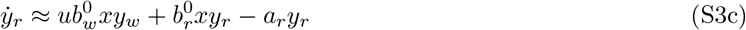

where, 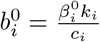.

At the infection equilibrium,

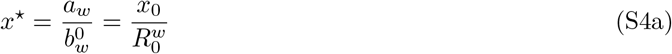

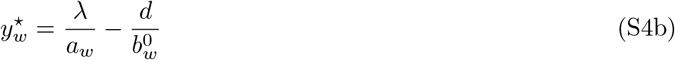

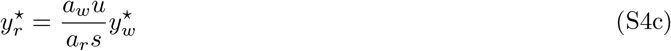

where 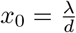 is the equilibrium level of target cells in the absence of infection. We assume for simplicity that the cost of resistance acts on viral infectivity, 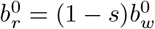 and all other infection parameters for the two strains are the same.

### Incorporating treatment

We model treatment by assuming that the drug blocks the infection of target cells. The infectivity parameter, *b*, becomes a function of the drug efficacy, *ϵ*

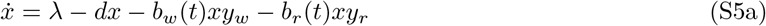

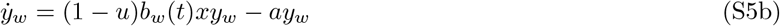

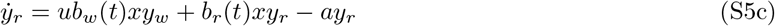

where 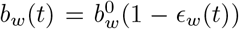 and 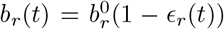. We are interested in the case when the WT is suppressed in the presence of treatment, *R*^*w*^ *<* 1 whereas the resistant strain is not *R*^*r*^ > 1.

**Figure S1** gives an example of what the dynamics look like when the treatment is initiated 100 days after the onset of infection. Prior to treatment, the system is at an equilibrium set by the WT. The resistant population is maintained at a low level via a mutation-selection balance. Once treatment starts, the WT is suppressed and the target cells start recovering. The resistant population decreases initially after treatment starts. This is on account of a lack of target cells to infect and a reduced influx from the suppressed WT population via mutation. The resistant population starts growing and establishes infection once the target cell availability is higher. Note that the resistant strain does not have to wait until the WT infection is fully cleared before it can start establishing infection; it can do so as soon as there are enough target cells around to infect.

A deterministic approach guarantees the establishment of a resistant infection once the WT is suppressed during treatment, so long as *R*^*r*^ > 1. However, mutations are inherently stochastic and the presence of just a few mutants prior to the start of treatment isn’t enough to guarantee establishment as they can undergo stochastic extinction. In order to calculate the establishment probability of a resistant infection emerging during treatment, we use a stochastic version of the model. It is a continuous time Markov Chain with the transition rates given by the rates in the differential equations [29].

### Modeling latency

To model latency as a source of resistance, we consider the scenario when the WT strain is completely suppressed by treatment and target cells have recovered to their pre-infection equilibrium 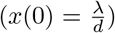. WT infected cells are introduced into the system via reactivation of latent infected cells and continue to be suppressed by treatment. We assume that prior to treatment the mutant was maintained at a mutation-selection balance [43, 80] which is reflected in the composition of the latent pool, a fraction 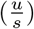 of the latent pool consists of cells infected by the resistant strain. This leads to the following system of equations for the target cells (*x*), WT (*y*_*w*_) and resistant mutant (*y*_*r*_) infected cells,

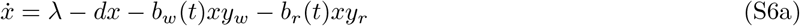

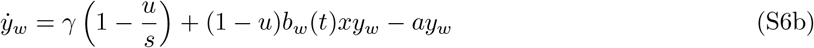

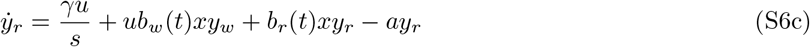

where *γ* is the rate of latency reactivation and all other parameters are the same as in **Equation S5**.

### Infection parameter values

**Table S1:** Infection parameter values used throughout this work unless stated otherwise.

| Parameter | Description | Units | Value |
| --- | --- | --- | --- |
| $\lambda$ | Rate of target cell production | cells/day | $2 \times 10^6$ |
| $d$ | Death rate of target cells | $\text{day}^{-1}$ | 0.1 |
| $k$ | Virion production rate | $\text{day}^{-1}$ | 1000 |
| $a$ | Death rate of infected cells | $\text{day}^{-1}$ | 0.5 |
| $c$ | Free virion clearance rate | $\text{day}^{-1}$ | 10 |
| $\beta$ | Viral infectivity | $\text{day}^{-1}$ | $5 \times 10^{-10}$ |
| $u$ | Mutation rate | | $3 \times 10^{-5}$ |
| $s$ | Cost of resistance | | 0.3 |

These parameters give a resulting *R*_0_ = 2 for the wild-type and *R*_0_ = 1.4 for the resistant mutant. We chose these parameter values to be consistent with Alexander and Bonhoeffer [29]. They provide their original sources and justify in detail why they are reasonable for the diseases of interest (for example HIV, HBV and HCV). We stress that these values are only meant to qualitatively capture the nature of the infection dynamics. Pathogen specific values should be used in practice while evaluating the risks associated with new therapies.

### Drug kinetics models

Throughout the paper we consider two example drug efficacy profiles with different relative rates of drug absorption and clearance. The first is a slow absorbing drug for which we use a simple sinusoidal function for the time-dependent drug efficacy (**Figure 1B**),

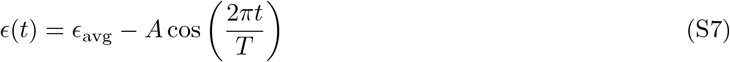

where, *ϵ*_avg_ is the time-averaged drug efficacy, *A* is difference between the average and peak or trough efficacy (amplitude), and *T* is the dosing interval (period). Although a heuristic function, we interpret this model as follows: When a dose is taken, the efficacy starts at the lowest level, and then increases as the drug is absorbed in the body and eventually reaches a maximum level (half-way through the dosing period). After this time, drug clearance dominants, lowering the concentration until the next dose is taken and the cycle repeats. The average drug efficacy may be different for WT and resistant strains.

Secondly, we consider a fast absorbing drug model where the efficacy is related to the drug concentration by a Hill dose-response curve (**Figure 1C**). We assume that drug absorption is rapid as compared to drug clearance (as is the case for many drugs), and approximate the drug concentration *D*(*t*) as an instantaneous jump to the peak level followed by exponential decay. Together, this gives

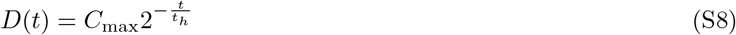

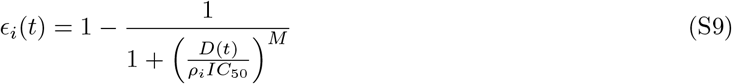

The maximum drug concentration is *C*_max_ and the half-life is *t*_*h*_. Lowest drug concentration (*C*_min_) for a drug with dosing period *T* occurs at the end of each dosing period that is, 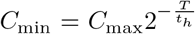. The drug concentration reaches *C*_max_ instantaneously after the first dose is taken at *t* = 0. Each dose increases the drug concentration by Δ*C* = *C*_max_ − *C*_min_. The efficacy of the drug against strain *i* at time *t* is *E*_*i*_(*t*), where, *IC*_50_ is the drug concentration that achieves half-maximal efficacy, *M* quantifies the steepness of the dose-response curve, and *ρ*_*i*_ corresponds to the extent of drug resistance for a particular strain (*ρ*_*i*_ = 1 for WT and *ρ*_*i*_ > 1 if resistant).

In the slow absorbing drug model, we vary the drug dosing period, *T*. In the fast absorbing drug model, we varied the drug half-life, *t*_*h*_ along with the dosing period.

We consider these two different models as existing long-acting drugs for other infections are known to have large differences in their absorption profiles [44]. There are currently only a few examples of long-acting antimicrobial therapies, since they are mainly in early stages of development, but we so far see examples of both slow and fast absorbing drugs. The only currently FDA-approved long-acting treatment for HIV - injectable cabotegravir (CAB) and rilpivirine (RPV) combination therapy - reaches a peak concentration before the middle of the dosing interval, with Tmax ∼ 1 week for a monthly dosing regimen [82] (example of a fast absorbing drug). For other long-acting therapies being considered for pre-exposure prophylaxis for HIV, drug concentrations peak closer to the middle of the dosing interval [83]. For example, once-monthly intramuscular injections of 800 mg CAB result in peak drug levels at around 2 weeks, while the same administration mode for 600 mg RPV results in a peak after 10 days (examples of slow absorbing drugs).

There are more examples for other drug classes with a longer history of extended-release formulations. Correll et al [44] provide a detailed review of the pharmacokinetics of long-acting antipsychotic drugs currently available to treat schizophrenia. They highlight the large differences in the absorption profiles across drugs; for example, aripiprazole lauroxil NanoCrystal dispersion 8 week dosing has time to peak Tmax ∼ 4 weeks, and paliperidone palmitate 4 week dosing has Tmax ∼ 2 weeks (e.g. two examples of slow absorbing drugs), while haloperidol decanoate 4 week dosing as Tmax ∼ 1 week, and olanzapine pamoate 4 week dosing as Tmax ∼ 4 days (e.g. two examples of fast absorbing drugs).

### Parameter values used for the drug kinetics models

In Figure 1 the parameter values used for the slow absorption model are *ϵ*_avg_ = 0.75, *A* = 0.20 and for the fast absorption model, *C*_max_ = 20, *IC*_50_ = 10, *M* = 10, *t*_*h*_ = *T* and *ρ* = 1 with drug concentrations in arbitrary units. In Figure 2 the parameter values used for the drug kinetics for the slow absorption model are *ϵ*_avg,w_ = 0.9, *A*_*w*_ = 0.09, *ϵ*_*r*_(*t*) = 0 (purple) and *ϵ*_avg,r_ = 0.1, *A*_*r*_ = 0.09 (green) and for the fast absorption model, *C*_max_ = 20, *IC*_50_ = 10, *M* = 10, 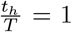, *ρ*_*w*_ = 1, *ρ*_*r*_ = ∞ (purple) and *ρ*_*r*_ = 2 (green). In Figures 3C,D the parameter values used for the drug kinetics, *C*_max_ = 20 (purple curve), *C*_max_ = 17 (green curve), *IC*_50_ = 10, *M* = 10, 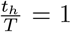, *ρ*_*w*_ = 1, *ρ*_*r*_ =∞ in C) and *ρ*_*r*_ = ∞ in D). In Figure 3E the parameter values used for the drug kinetics, *ϵ*_avg,w_ = 0.9, *A*_*w*_ = 0.09 (purple curve) and *ϵ*_avg,w_ = 0.6, *A*_*w*_ = 0.25 (green curve). The purple curve corresponds to a high drug efficacy such that *R*^*w*^ *<* 1 during all points of the drug cycle. *R*^*w*^ > 1 for ∼ 30% of each period for the low efficacy drug cycle given by the green curve. Figures 4A,B,D and E have the same parameter values as the fast absorbing drug from Figure 2 and Figures 4C and F have the same parameter values as in Figure 3.

### Establishment probability

We calculate the probability that a resistant mutant existing at a certain time after treatment initiation avoids extinction by modeling the process as a time inhomogeneous “Birth-Death” (B-D) process [84]. From **Equation S5c** births and deaths occur at the per capita rate,

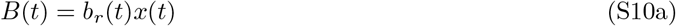

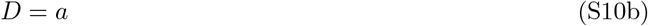

At the beginning of treatment, target and WT infected cells are present in very large numbers in contrast to the resistant mutant. So, we approximate that the target and WT infected cells follow deterministic dynamics given by the following simplified system of equations which can be solved numerically (we use Mathematica 12.0 [85]),

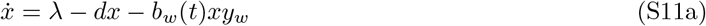

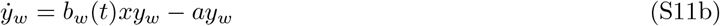

There exists an analytical solution for the extinction probability of such a B-D process [84],

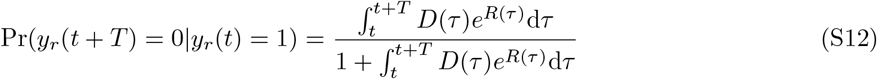

where 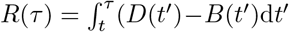. The probability that the lineage of a strain existing at time *t* eventually goes extinct is given by the limit,

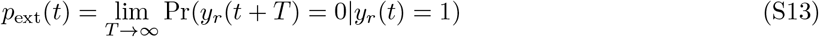

the establishment probability is,

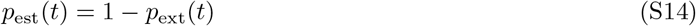

This “Birth-Death” approximation is used instead of stochastically simulating the whole model for each dosing interval as it captures the effects of stochasticity well and has the added advantage of calculating the risks significantly faster, within minutes as opposed to days. The approximation works well due to the deterministic nature of the target and WT infected cell dynamics coupled with the small population size of the resistant mutant at the onset of treatment. It does however have a limitation as it doesn’t include the depletion of the target cells by the resistant strain (**Equation S11**). As a result, the establishment probability isn’t well approximated when there is rapid growth of the resistant strain early on after treatment is initiated. But, this only affects the numerical results and doesn’t change the qualitative trends associated with the risk of resistance and drug dosing intervals and the method can still be used to assess the relative risks associated with longer-acting therapies.

### Different sources of resistance

Drug resistance can arise via different sources: mutants existing prior to treatment (pre-existing), those that are generated de novo during treatment via residual wild-type infection (rescue) and those that are a result of latency reactivation.

### Pre-existence

The establishment probability for a single mutant present at the start of treatment is given by **Equation S14** with *t* = 0.

It is also possible to derive the total probability of survival due to pre-existence by taking into account the full distribution of the mutant population size prior to treatment. This can be approximated by a steady state birth-death-immigration (BDI) process [29]. The target and WT infected cell populations are large enough that fluctuations in their size aren’t important and we can assume they are at their deterministic equilibrium level. This leads to a BDI process with constant rates where “birth” refers to the replication of a mutant infected cell; “death” is the death of a mutant infected cell; and “immigration” is the continual flow into the mutant population due to de novo mutations in the WT. We can read off these rates from **Equation S3c** substituting the values of *x*^⋆^ and 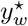 from **Equations S4a and S4b** to give,

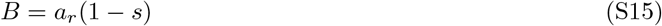

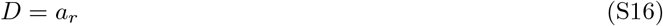

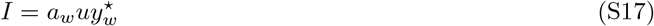

The probability generating function (PGF) for the steady state distribution when *B < D* is given by [84],

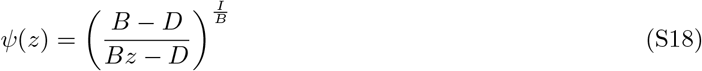

With the mean 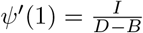. The total survival probability is given by,

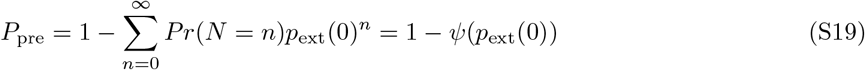

### Rescue

The rate of appearance of resistant mutants is proportional to the level of residual infection of the WT and the mutation probability. This rate can be read off from **Equation S4c**,

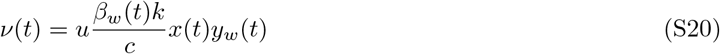

Once a mutant is produced at time *t*′, its establishment probability is given by **Equation S14** calculated at *t* = *t*′. Since we want to keep track of only those mutants that avoid extinction, the total rate of producing rescue mutants is,

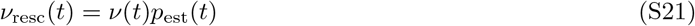

We model the number of rescue events that occur via a time inhomogeneous Poisson process [29] with the above rate and calculate probability that at least one rescue mutant establishes infection,

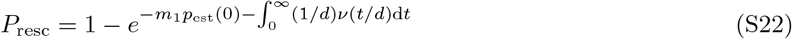

where 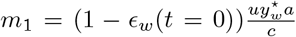 is a correction term that is needed because of the quasi-equilibrium approximation used for setting the free virus population proportional to the infected cells [29].

### Latency reactivation

For mutants produced via latency reactivation, the establishment probability *p*_est_(*t*) is calculated as before by modeling this as a Birth-Death process with rates, *B*(*t*) = *b*_*r*_(*t*)*x*(*t*) and *D* = *a*. We approximate the target and WT infected cell dynamics by solving the following deterministic system,

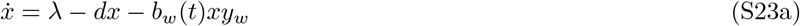

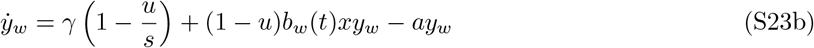

The rate at which resistant mutants are produced due to latency reactivation can be read off from **Equation S6**,

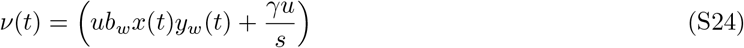

In this case we also just keep track of those mutants that escape stochastic extinction and the total rate is given by,

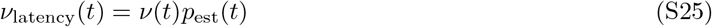

and the average probability of rescue per latency reactivation event is,

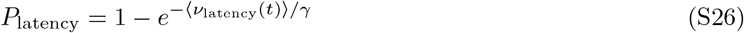

where, ⟨*ν*_latency_(*t*)⟩ is the rate averaged over one period of the drug cycle. The establishment of resistance due all the three sources was computed numerically using Mathematica 12.0 [85].

### Stochastic simulations of the birth-death approximation

We checked the validity of the birth-death approximation used for all the three sources of resistance. For pre-existence and rescue, we simulated the full model (**Equation 1**) stochastically using Gillespie’s tau-leaping method [86]. The step size, Δ*t* was fixed to 0.001 time units. In this algorithm, the number of each type of event occurring in one time-step is drawn from a Poisson distribution with a mean = event rate × Δ*t*. To obtain the establishment probability of a single pre-existing resistant mutant, we simulated the treatment phase multiple times (10, 000 − 100, 000 iterations). The initial conditions for the target cells and the WT infected cells were given by their pre-treatment equilibrium values with one resistant infected cell. We set the mutation rate to zero to isolate the establishment due to just the pre-existing mutant. The simulations were run long enough such that the mutant population had either established infection or gone extinct. The fraction of runs with surviving mutants estimates the establishment probability. For the rescue probability, we ran multiple stochastic simulations of the full model (10, 000 iterations) with the same pre-treatment initial conditions for the target and WT infected cells. We set the initial resistant infected cell population to zero and included a non-zero mutation rate. The simulations were run long enough such that either a resistant infection had been established or the infection was cleared. The rescue probability corresponds to the fraction of runs in which the resistant infection was established.

To obtain the average probability of rescue per latency reactivation event, we stochastically simulated **Equation S6** in three stages using Gillespie’s tau-leaping method with the step size Δ*t* = 0.001. At the start of the simulation, the target cells are at their pre-infection equilibrium with no WT or resistant infected cells. In the first stage of the simulation we turn off the mutation rate and allow the WT population to settle into a form of an equilibrium where suppression by treatment is balanced by the activation of latent cells. In the second stage of the simulation we allow the reactivation of resistant latent cells and turn on the mutation rate for a period of 1*/γ*. Latency reactivation is a Poisson point process with a constant rate *γ* where on average one reactivation event occurs during a period of 1*/γ*. This allows us to simulate the scenario of one reactivation event potentially leading to resistance. The probability of rescue (averaged over simulations) obtained in this manner is a good approximation of the probability per reactivation event as long as simulations where more than one reactivation event occurs do not contribute disproportionately to it. The probability of a resistant infection establishing due to *n* reactivation events, each of which has a probability *p* of causing infection, can be calculated by using the binomial distribution. In the limit of very small *p* (*<<* 1) however, this probability is well approximated by *np*. As we are in this small *p* regime, each reactivation event contributes equally to the probability and our approximation is valid. In the last stage, we turn off the mutation rate again and run the simulation for long enough such that the mutant has either gone extinct or established infection. Some of the probabilities involved are very small *O*(10^−5^) and in order to avoid running the large number of iterations, ∼ *O*(107) required to calculate these numbers (corresponding to weeks of computing time), we scaled up the mutation rate for the simulations and scaled down the final probability that was obtained. This is a reasonable step to take as when the probability is *<<* 1, it is directly proportional to the mutation rate. For each set of parameter values, we allowed the second stage of the simulation to occur at 100 different time-points in the drug cycle and repeated this for 200 iterations each. The final probability is given by the fraction of runs (20, 000 in total) where the mutant established infection.

All the simulations were implemented in Python 3.0 and run on Cannon, the Harvard University cluster.

### Existence of the establishment time-frame

In order to demonstrate the dependence of pre-existence and rescue probabilities on a short initial time-window after treatment onset, we computed the fraction of pre-existing mutants that have survived as a function of time after treatment onset (**Figure S2**). This can also be interpreted as the probability that the lineage of one pre-existing strain has avoided extinction by time *t, P* (*y*_*r*_(*t*) ≠ 0|*y*_*r*_(0) = 1). This probability is bounded by *P* (*y*_*r*_(0) ≠ 0|*y*_*r*_(0) = 1) = 1 and *P* (*y*_*r*_(∞)≠ 0|*y*_*r*_(0) = 1) = *p*_est_. If the establishment probability is reached in a finite amount of time, this implies that the lineage is guaranteed to establish infection if it survives until then. We find that this occurs rather quickly after treatment starts, and is inversely related to the infected cell decay rate *a*. Given the non-linear nature of the infection and treatment dynamics it is difficult to precisely measure the establishment time-frame. However, we don’t consider this to be a limitation of our analysis as it is not needed to calculate the risks of resistance. The usefulness of the concept lies in helping understand and interpret the trends seen on varying drug dosing intervals on the risk of resistance.

### Models of non-adherence

#### Random adherence model

In the random adherence model we assume that there is a constant probability of missing each dose apart from the first one which is always taken. The establishment and rescue probabilities were computed numerically using Mathematica 12.0 for the fast-absorbing (pharmacologically-inspired) efficacy model.

#### Delayed adherence model

We consider an alternative adherence model to capture the scenario where an individual on long-acting therapy may take a missed dose on any subsequent day instead of waiting until the next scheduled dose. To model this, we assume that there are two probabilities: a probability of missing the scheduled dose *p*_1_, and another non-adherence probability *p*_2_ such that on each following day the missed dose can be taken with probability 1 − *p*_2_. Once a dose is taken, the next dose is scheduled for *T* (the original dose period) days after. Consequently, the average non-adherence probability is given by,

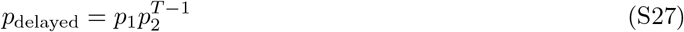

In order to get a desired *p*_delayed_, we fix *p*_1_ and calculate *p*_2_ for each dosing period. Note that here *p*_1_ has to be greater than *p*_delayed_. We chose *p*_1_*/p*_delayed_ = 1.5 so that the chance of missing a scheduled dose isn’t too high while ensuring that *p*_2_ doesn’t approach 1 too quickly for large *T*. For example, for an overall adherence probability of 70% the probability of missing the scheduled dose is *p*_1_ = 45%, and of missing each subsequent day is *p*_2_ = 67% for dosing every 2 days, *p*_2_ = 93% for weekly dosing, and *p*_2_ = 98.5% for monthly (4 week) dosing.

### Multi-step resistance pathways

The results in the main text are obtained from a model with only two strains (WT and a single resistant mutant), where one effective mutation rate encapsulates the effects of all the genetic changes needed to acquire drug resistance. In reality, resistance is often attained via step-wise accumulation of mutations [87, 88]. Furthermore, a spectrum of these pathogen variants may already exist prior to treatment initiation and can lead to a rapid emergence of drug resistance after the onset of therapy [89]. We hypothesized that the change in resistance risk due to long acting therapy may depend on the mutational pathway, since long-term exposure to sub-optimal drug levels can favor selection of strains of intermediate resistance. In order to investigate this, we extended the model to include an intermediate strain one mutational step away from the WT, and a resistant strain that is one further step away. In the absence of treatment, we assume that the WT is the fittest strain (**Figure S8A**), and with treatment the WT and the intermediate strain is suppressed but the resistant strain is not (**Figure S8A**).

#### Two-step mutations

##### Pre-existence

The establishment probability of one pre-existing mutant is independent of the mutational pathway to resistance from the WT strain but the frequency distribution of the resistant strain prior to treatment changes. We generalize the PGF for the point mutants given by **Equation S18** to two-step mutants using the method illustrated by Moreno-Gamez et al. [64] in their Supplementary Information.

An ODE model that can describe the dynamics of a system with a two-step mutant is given by [80, 91],

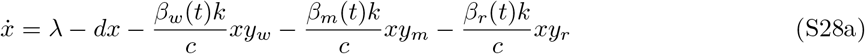

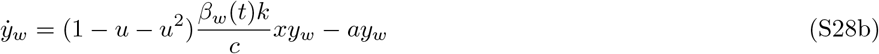

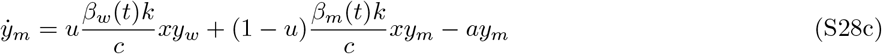

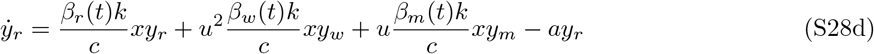

where, *y*_*m*_ is the intermediate strain a point mutation away from the WT with *u* as the mutation rate and *y*_*r*_ is the two-step mutant. The frequency distribution of the two-step mutant at the start of treatment can be approximated by considering the stochastic process determining the size of the single and double mutant populations (*y*_*m*_, *y*_*r*_). We make the assumption that the target cell and WT populations are at their deterministic pre-treatment equilibrium values (**Equations S4a and S4b**). The selection coefficients are assumed to be larger than the mutation rates to get the following approximate rates,

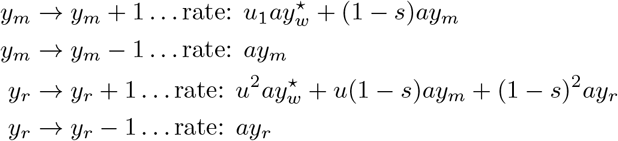

where *s* is the two mutation cost. At this point we make a further approximation as this is no longer a B-D-I process and doesn’t possess an analytic solution [64]. We make the approximation that if double mutants are frequent enough to affect treatment failure, then for realistic values of *u* and *s*, the single mutant should be frequent and well approximated by its constant equilibrium levels, 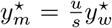. The stochastic system is modified to,

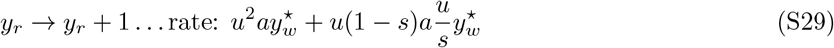

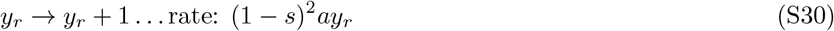

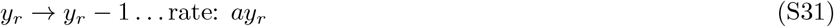

This is a modified B-D-I process with,

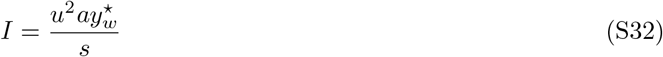

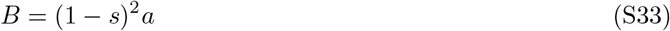

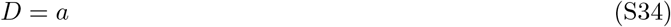

and the PGF can be calculated by **Equation S18**.

While pre-existing resistance mutations may be less frequent if they require multiple mutational steps from the wild type, the establishment probability of each mutant once treatment begins is independent of how it was produced. Consequently, the effect of dose frequency on the risk of resistance is the same as for single vs multi-step resistance pathways (**Figure 2**).

##### Rescue mutants

In contrast to pre-existence, the risk of resistance due to mutants produced during treatment (rescue) depends upon the rate of mutant production from the wild-type and the survival of intermediate strains, and consequently may depend upon the mutational pathway. As the establishment probability for one pre-existing mutant is unaffected by the mutational pathway to resistance, the only thing that changes in the case of the rescue mutants is their rate of production from the wild-type and intermediate strains.

**Equation S20** for the rate of resistant mutant production is modified to,

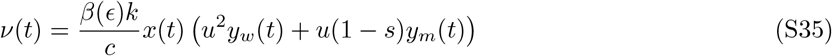

We analyzed the rescue probability of a partially-resistant mutant as a function of the drug dosing intervals for the fast-absorbed drug model for perfect and imperfect adherence. The choice of the drug kinetics model is only for illustrative purposes, we found that the main conclusion of this analysis did not depend on the drug absorption rates. We find that the trends are the same as that for one-step mutants, with long-acting therapies corresponding to a lower risk of resistance (**Figure S8B**). As long as the intermediate strain is suppressed by treatment, long-acting drugs that tend to stay near their peak efficacy during the important time-frame of establishment lead to lower rates of resistant mutant production, and in turn reduce the rescue probability.

We also calculated the risk of resistance from reactivation of persistent infection (“rescue” from reactivation) for a two-step mutant for both perfect (**Figure S8C**) and imperfect adherence (**Figure S8D**). Again, the trends are the same as that for one-step mutants but with a larger relative increase in resistance risks associated with long-acting drugs for treatments that aren’t fully suppressive. This is on account of an increase in the rate of resistant mutant production from the intermediate strains when exposed to the non-suppressive part of the drug cycle for longer.

Overall, we find that the mutational pathway considered here doesn’t change the qualitative relationship between the frequency of drug dosing and the risk of resistance that exists for the simplified case of a one-step mutant.

## Supplementary Figures

**Figure S1:**
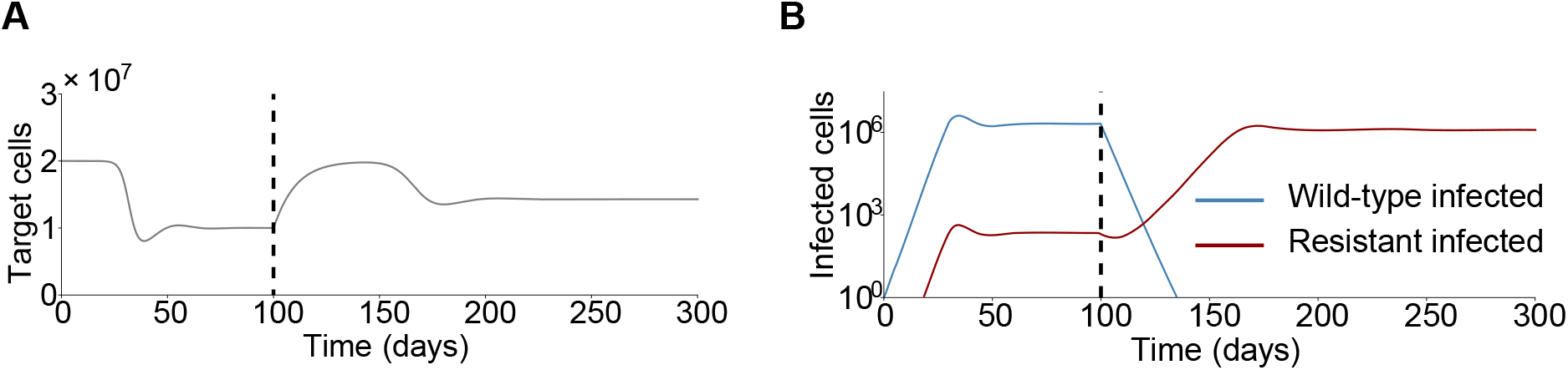
Example time-course of a chronic infection followed by treatment under the deterministic viral dynamics model. A) Target (uninfected) cells as a function of time. B) Time-course of cells infected by the wild-type (blue) and resistant (red) strain. Treatment is started *t* = 100 days (vertical dotted line) after the onset of infection. Parameter values, *λ* = 2 × 10^6^ cells*/*day, *d* = 0.1day^*−*1^, *k* = 1000day^*−*1^, *a* = 0.5day^*−*1^, *c* = 10day^*−*1^, *β*^0^ = 5 × 10^*−*10^day^*−*1^, *u* = 3 × 10^*−*5^ and *s* = 0.3. Initial conditions, 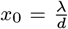, *y*_*s*0_ = 1 and *y*_*r*0_ = 0. Before treatment, *β*_*w*_ = *β*^0^, *β*_*r*_ = *β*^0^(1 − *s*). After treatment, *β*_*w*_ = *β* (1 − *ϵ*_*w*_) with *ϵ*_*w*_ = 0.9, *A*_*w*_ = 0 and *ϵ*_*r*_ (*t*) = 0. For these values of the average drug efficacy and viral dynamics parameters, the WT infection is cured during the first treatment cycle for drug dosing intervals roughly greater than 5 weeks.

**Figure S2:**
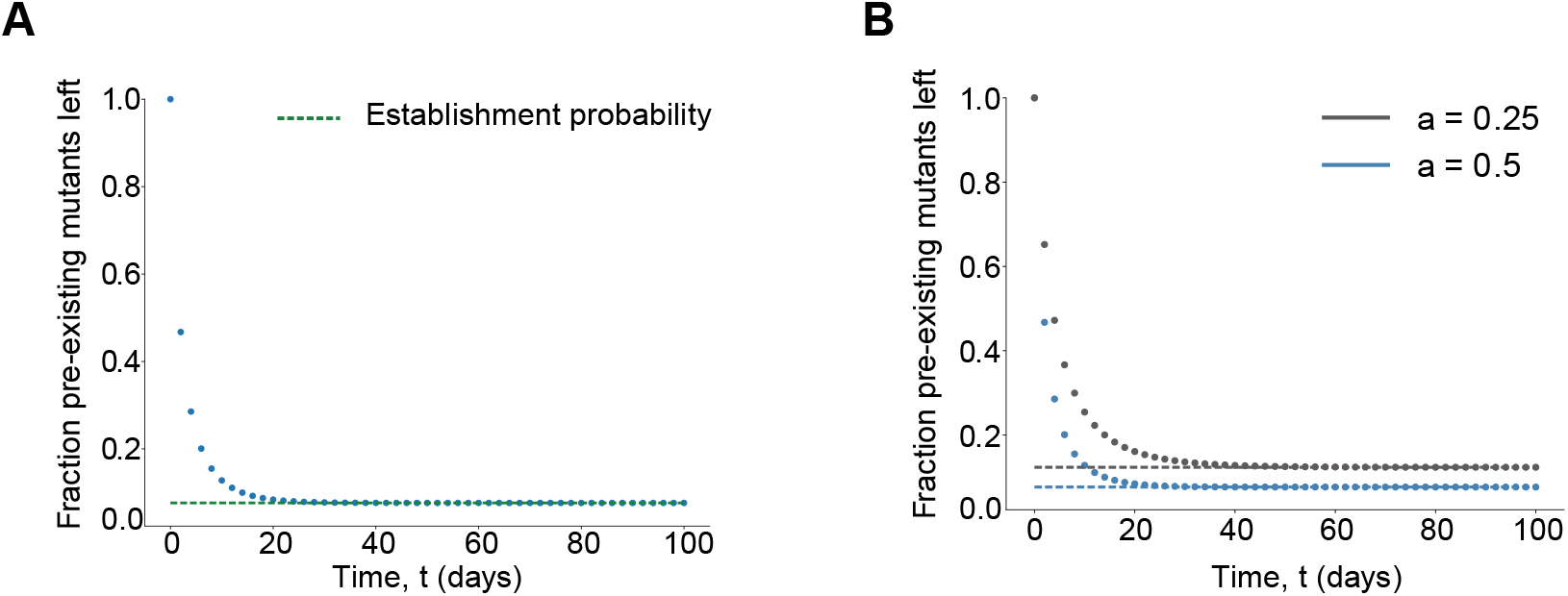
Fraction of pre-existing mutants left as a function of time post treatment onset. The dotted lines correspond to the establishment probability for a fully-resistant mutant in the presence of the fast absorbing drug model with dosing period T = 1 with A) infected cell decay rate *a*=0.5 and B) with two values of the infected cell decay rate *a* = 0.5(green) and *a* = 0.25(purple). The burst rate *k* is adjusted according to the value of *a* to ensure that *R*_0_ for the two strains is the same in both cases. All other parameter values are the same as in Figure 2B in the main text.

**Figure S3:**
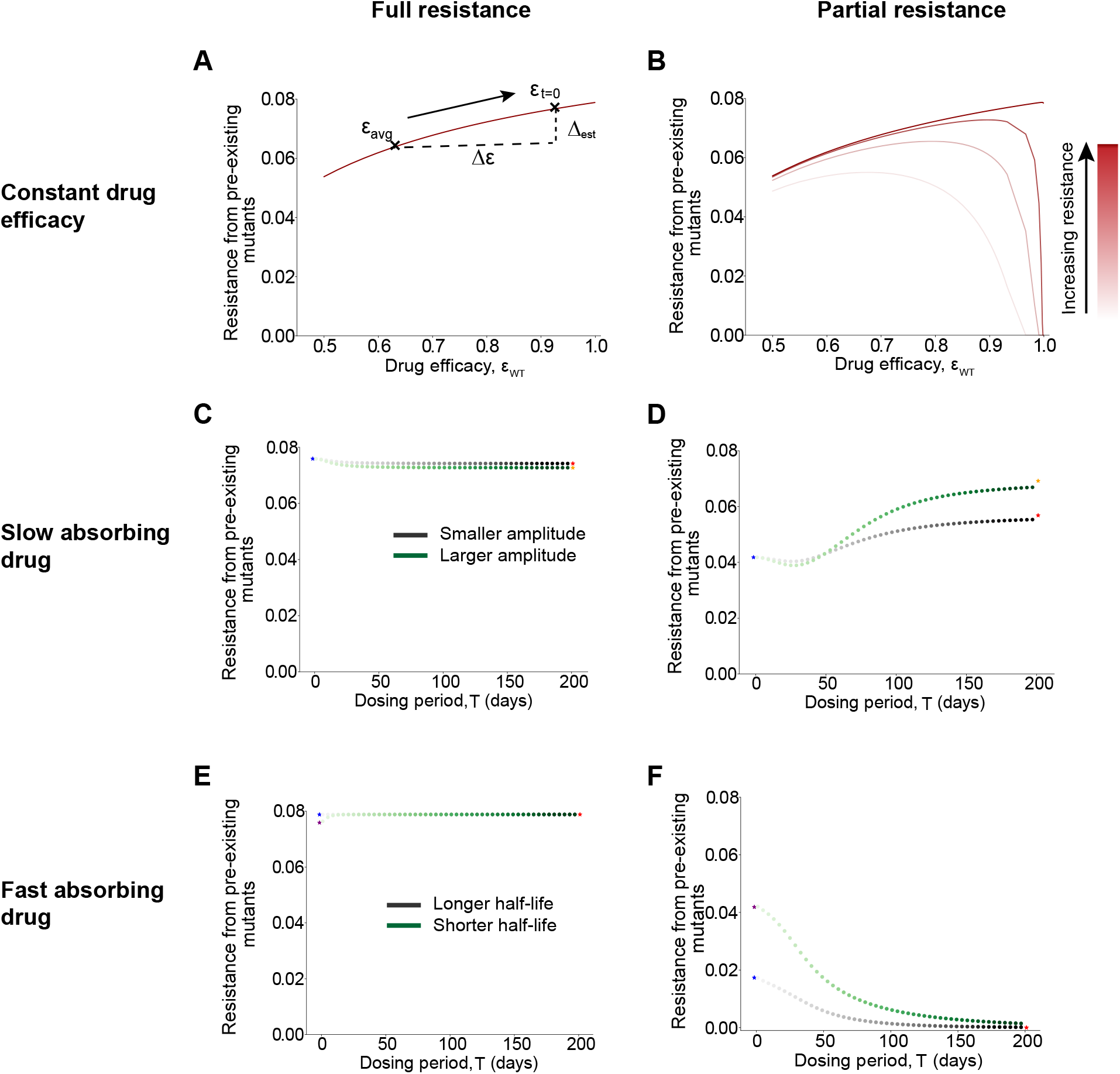
Effect of varying drug efficacy parameters on the establishment probability of pre-existing mutants. A)-B) Establishment probability as a function of constant drug efficacy for A) fully resistant (*ρ*_*r*_ = ∞) B) partially resistant mutants (*ρ*_*r*_ = 1.5, 1.7, 2, 4 with increasing darkness). Black marks in A) are for illustrative purposes to show the effect of increasing the drug dosing interval on the establishment probability for a fast absorbing drug (*ϵ*(*t* = 0) *> ϵ*_avg_). Δ_est_ and Δ*ϵ* are the change in establishment probability and drug efficacy respectively. See Supplementary Discussion for more details. C)-D) Effect of varying the amplitude keeping the average efficacy fixed in the slow absorbing drug model for a C) fully resistant and D) partially resistant mutant (*ρ*_*r*_ = 2). Parameter values for the green curves are the same as in Figure 2 in the main text with *A* = 0.05 for the grey curves. The starred points correspond to the establishment probability for constant efficacies, time-averaged efficacy (blue stars) and efficacy at *t* = 0 (orange and red stars) E)-F) Effect of varying the half-life *t*_*h*_ in the fast absorbing drug model keeping *C*_max_ fixed for a E) fully resistant and F) partially resistant mutant (*ρ*_*r*_ = 2). Parameter values for the green curves are the same as in Figure 2 in the main text with *t*_*h*_*/T* = 5 for the grey curves. The starred points correspond to the establishment probability for constant efficacies, time-averaged efficacy (blue and purple stars) and efficacy at *t* = 0 (red stars).

**Figure S4:**
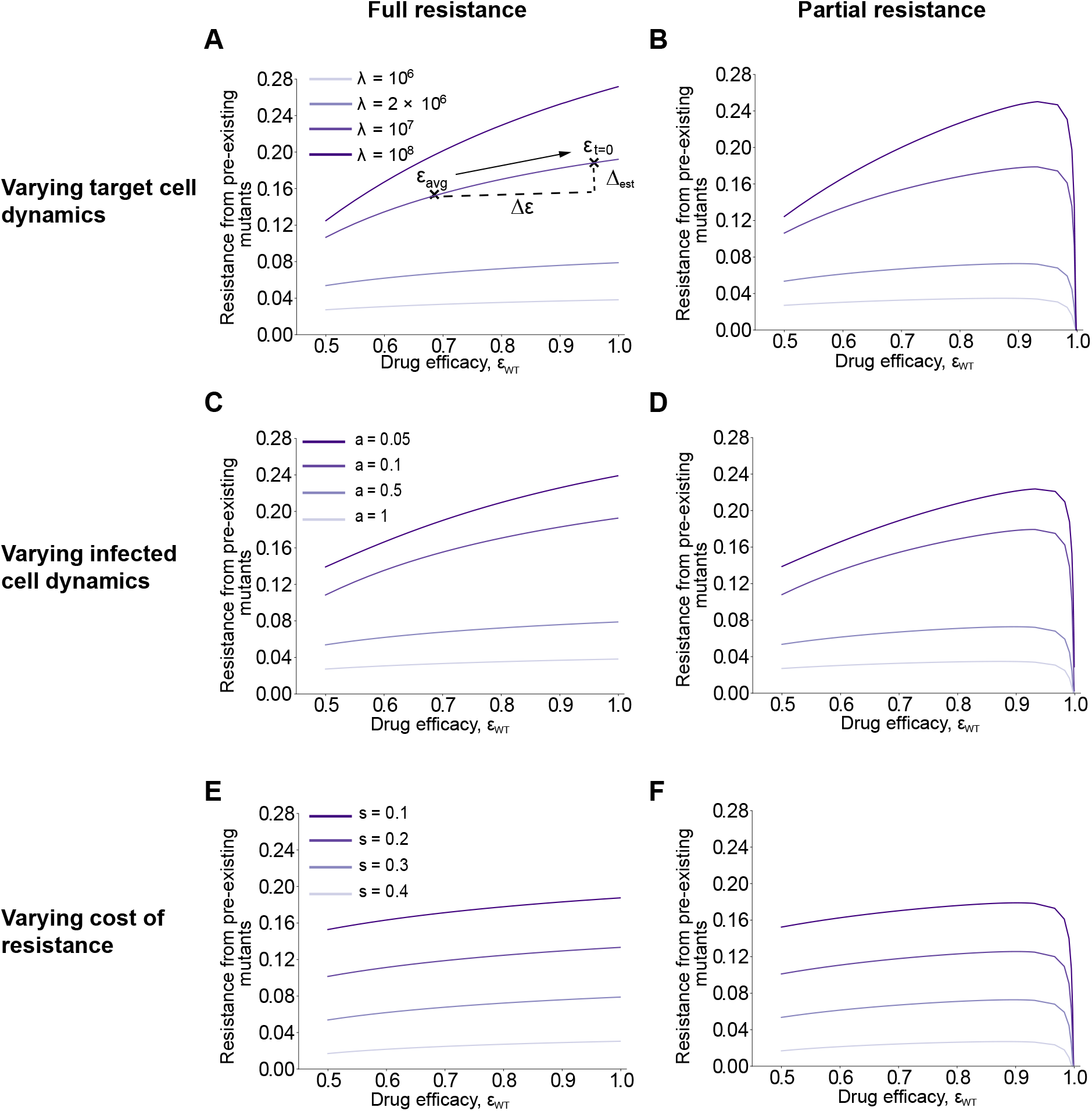
Effect of varying infection parameters on the establishment probability of pre-existing mutants in the presence of constant drug efficacy. A)-B) Establishment probability on varying the target (uninfected) cell production rate *λ* for a A) fully and B) partially resistant (*ρ*_*r*_ = 2) mutant. *λ* increases as the color darkens with the death rate of uninfected cells *d* increasing proportionally to keep *R*_0_ for the wild-type fixed. Black marks in A) are for illustrative purposes to show the effect of increasing the drug dosing interval on the establishment probability for a fast absorbing drug (*ϵ*(*t* = 0) *> E*_avg_). Δ_est_ and Δ*ϵ* are the change in establishment probability and drug efficacy respectively. See Supplementary Discussion for more details. C)-D) Establishment probability for a C) fully and D) partially resistant (*ρ*_*r*_ = 2) mutant on varying the infected cell death rate *a. R*_0_ for the wild-type is kept fixed by varying the infectivity parameter *b* proportionally with *a*. E)-F) Effect of varying the cost of resistance *s* for a E) fully and F) partially resistant (*ρ*_*r*_ = 2) mutant on the establishment probability. *s* decreases as the color darkens. The efficacy on the x-axis corresponds to the wild-type drug efficacy. All other parameter values are the same as in Figure S1.

**Figure S5:**
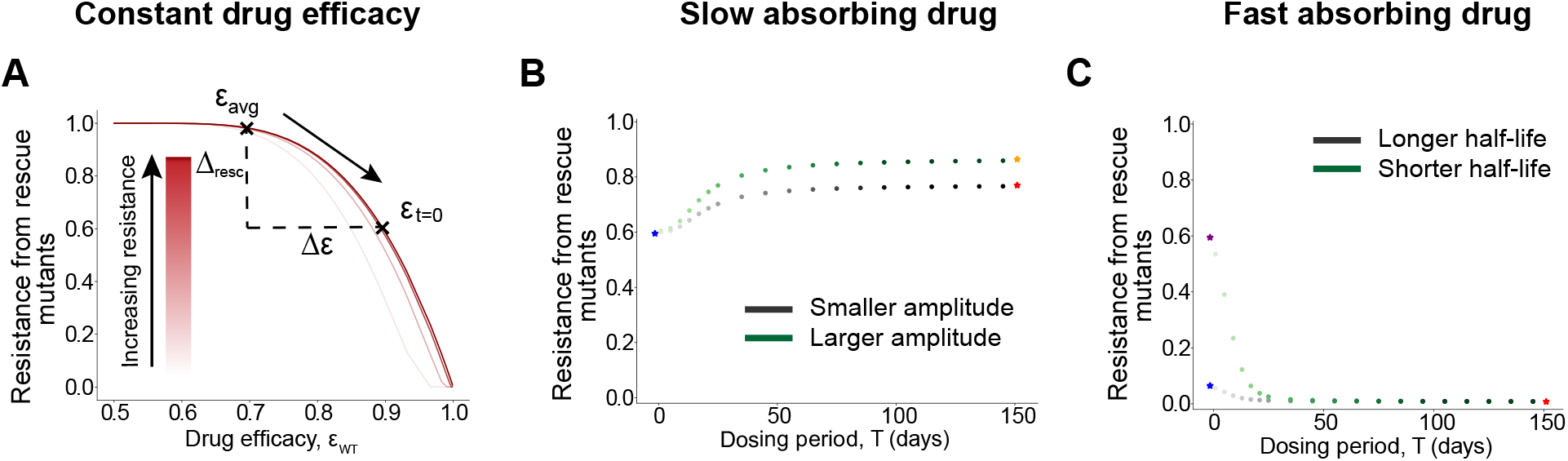
Effect of varying drug efficacy parameters on the rescue probability. A) Rescue probability as a function of constant efficacy for fully (*ρ*_*r*_ = ∞) and partially resistant (*ρ*_*r*_ = 1.5, 1.7, 2, 4 with increasing darkness) mutants. Black marks in A) are for illustrative purposes to show the effect of increasing the drug dosing interval on the rescue probability for a fast absorbing drug (*ϵ*(*t* = 0) *> ϵ*_avg_). Δ_resc_ and Δ*ϵ* are the change in rescue probability and drug efficacy respectively. See Supplementary Discussion for more details. B) Effect of varying the amplitude keeping the time-averaged drug efficacy fixed in the slow absorbing drug model for a fully resistant mutant. Parameter values for the green curves are the same as in Figure 2 in the main text with *A*_1_ = 0.05 for the grey curves. The starred points correspond to the rescue probability for constant efficacies, time averaged efficacy (blue stars) and efficacy at *t* = 0 (orange and red stars) C) Effect of varying the half-life *t*_*h*_ in the fast absorbing drug model keeping *C*_max_ fixed for a fully resistant mutant. Parameter values for the green curves are the same as in Figure 2 in the main text with *t*_*h*_*/T* = 5 for the grey curves. The starred points correspond to the rescue probability for constant efficacies, time averaged efficacy (blue and purple stars) and efficacy at *t* = 0 (red star).

**Figure S6:**
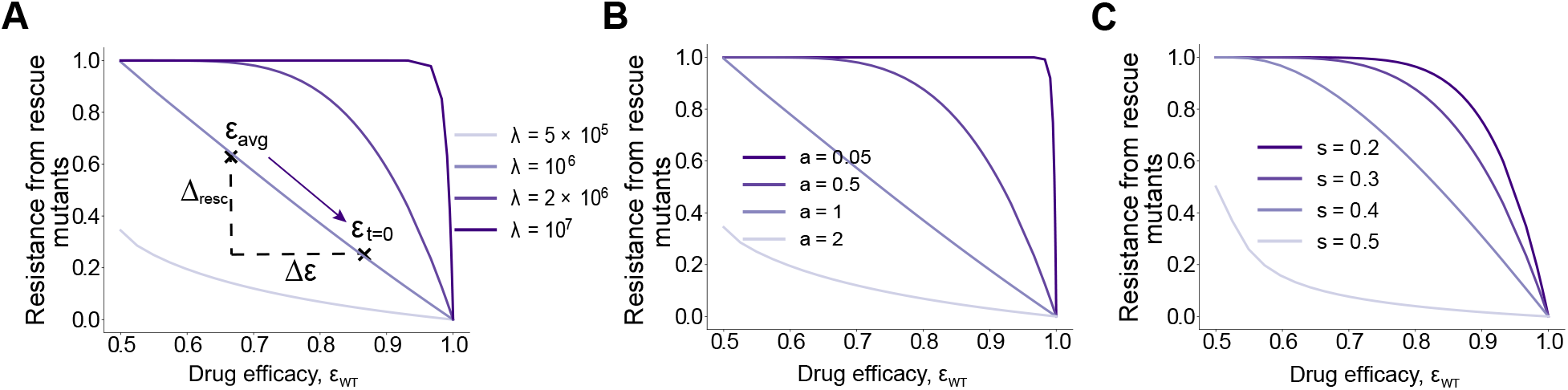
Effect of varying infection parameters on the rescue probability. A) Rescue probability on varying the target (uninfected) cell production rate *λ* for a fully resistant mutant as a function of a constant drug efficacy. *λ* increases as the color darkens, with the death rate of uninfected cells *d* increasing proportionally to keep *R*_0_ of the wild-type fixed. Black marks in A) are for illustrative purposes to show the effect of increasing the drug dosing interval on the rescue probability for a fast absorbing drug (*ϵ*(*t* = 0) *> ϵ*_avg_). Δ_resc_ and Δ*ϵ* are the change in rescue probability and drug efficacy respectively. See Supplementary Discussion for more details. B) Effect of varying the infected cell death rate *a* on the rescue probability for a fully resistant mutant as a function of a constant drug efficacy. *a* decreases as the color darkens. C) Rescue probability on varying the cost of mutation *s* for a fully resistant mutant as a function of a constant drug efficacy. *s* decreases as the color darkens.

**Figure S7:**
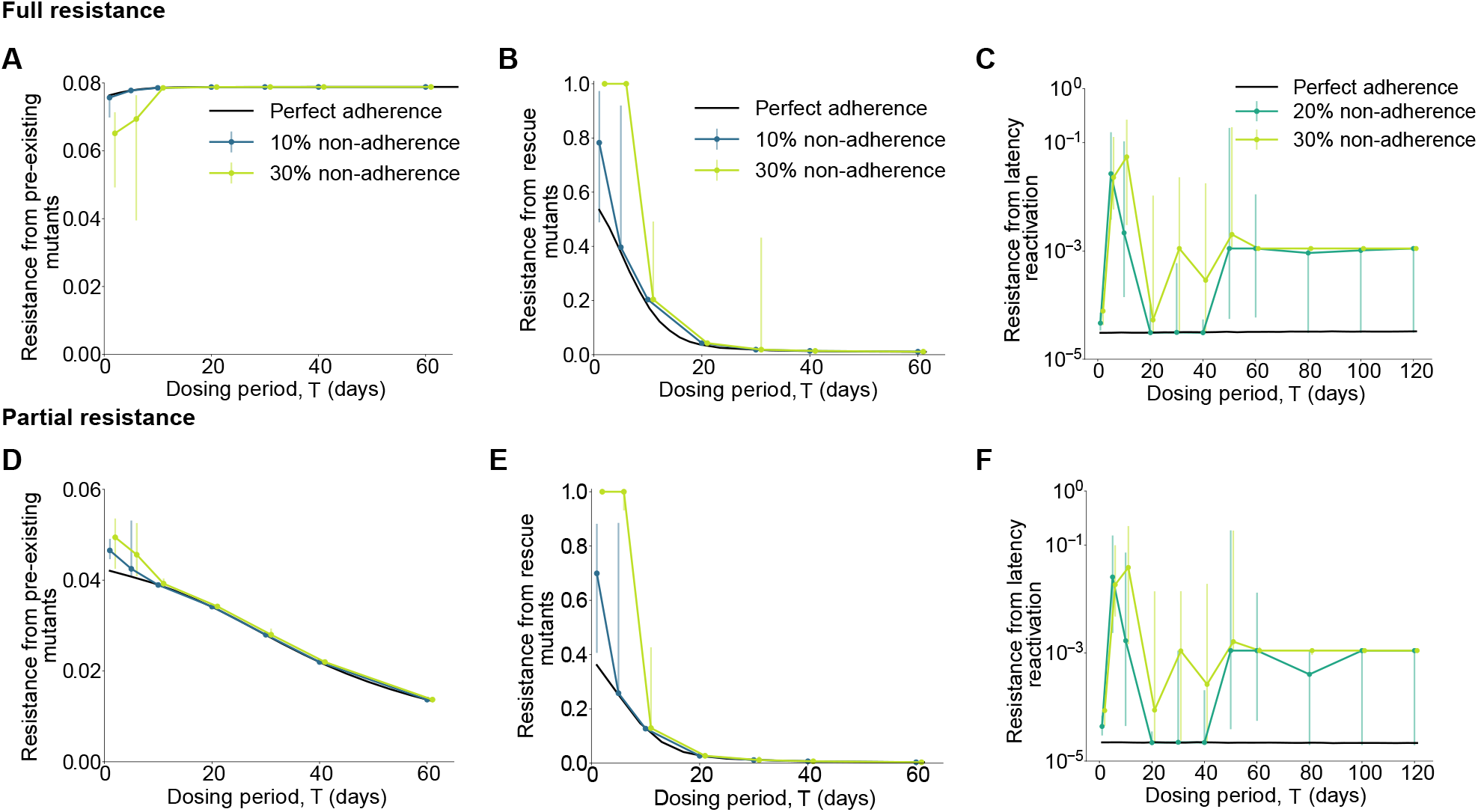
Effect of non-adherence on the establishment of resistance for different drug dosing intervals for a delayed adherence model. Establishment probability of A) fully (*ρ*_*r*_ = ∞) and D) partially (*ρ*_*r*_ = 2) resistant pre-existing mutants. Probability that at least one B) fully (*ρ*_*r*_ = ∞) and E) partially (*ρ*_*r*_ = 2) resistant rescue mutant is produced. Average probability of rescue per latency reactivation event for a C) fully (*ρ*_*r*_ = ∞) and F) partially (*ρ*_*r*_ = 2) resistant mutant. These results are under the fast absorbing drug model with the solid black line corresponding to results for perfect adherence. The average probability of non-adherence in this model is given by 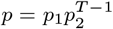 where *p*_1_ is the probability of missing a scheduled dose, 1 − *p*_2_ is the daily probability with which the dose is taken on subsequent days, and *T* is the dosing interval. For *p* = 0.1, 0.2, 0.3 we fix *p*_1_ = 0.15, 0.3, 0.45 and calculate the value for *p*_2_ based on the value of the dosing interval. Imperfect adherence results are medians of 100 iterations for pre-existence and rescue, and 500 iterations for latency. The error bars correspond to the interquartile range. X-axis positions have been offset for ease of visualization.

**Figure S8:**
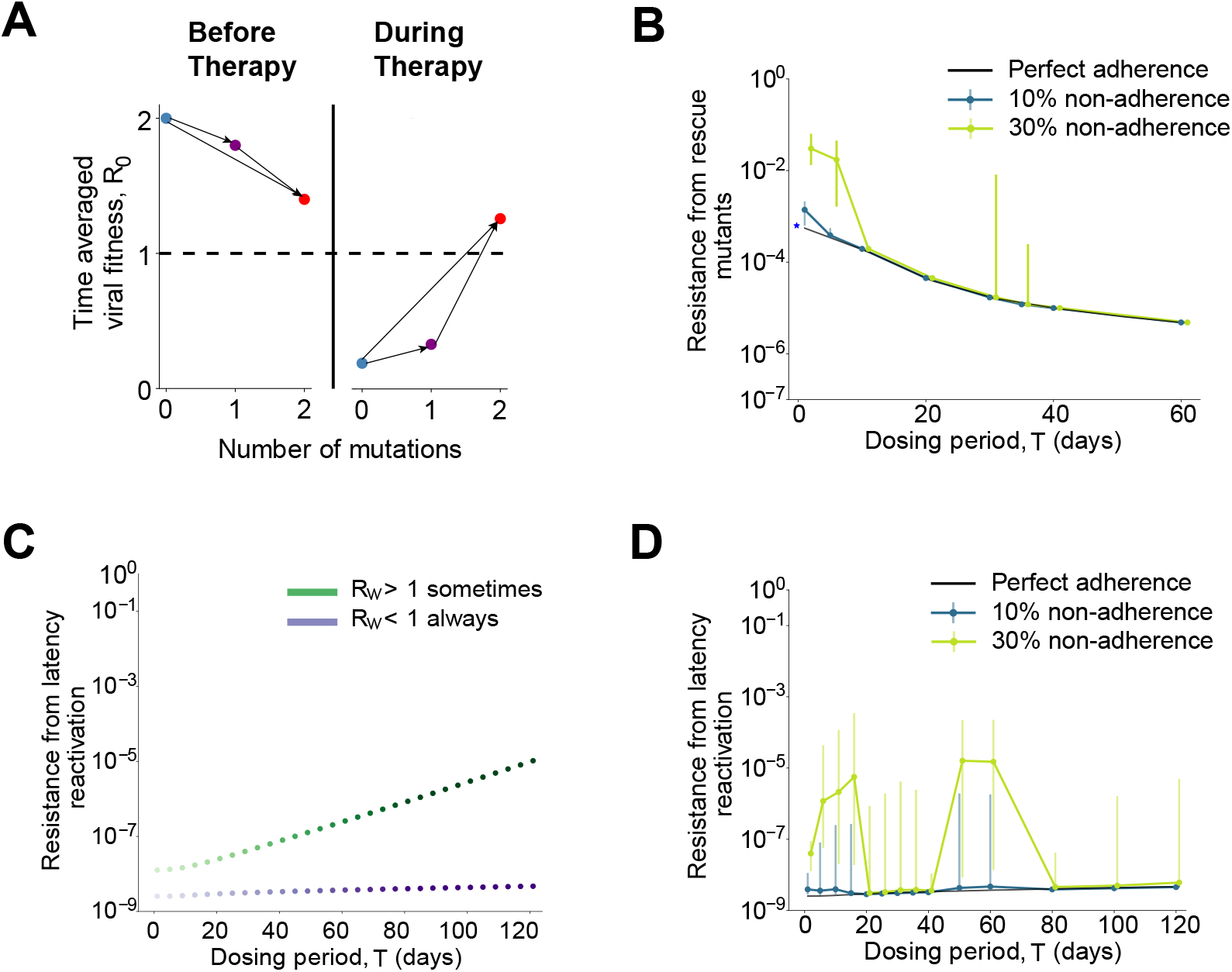
Effect of drug dosing intervals on the establishment of resistance for two-step mutants. Results are for a two-step partially resistant mutant and fast absorbing efficacy model. A) Time-averaged viral fitness of the WT (blue dot), intermediate strains (purple dot) and resistant strain (red dot), before and during treatment. B) Probability that at least one rescue mutant is produced as a function of the dosing interval for perfect (black line) and imperfect adherence. The blue starred point corresponds to the rescue probability for constant efficacy (blue star). C) Average probability of rescue per latency reactivation event as a function of the dosing interval for two strengths of the efficacy profile with perfect adherence. For the green curve, *R*_0_ for the WT > 1 for ∼ 23% of the cycle and > 1 for ∼ 35% of the cycle for the intermediate mutants. D) Effect of imperfect adherence on the average probability of rescue per latency reactivation event. The black line corresponds to perfect adherence for the strong drug efficacy from C). Results are medians of 100 iterations for rescue and 500 iterations for latency with the error bars corresponding to the interquartile range.

## Supplementary Discussion

### Role played by treatment parameters on the establishment probability for pre-existing mutants

#### Mutants fully resistant to treatment

In the presence of a constant drug efficacy, the establishment probability for mutants fully resistant to treatment is higher as the drug efficacy increases (**Figure S3A**). Stronger wild-type (WT) suppression is beneficial to the mutant strain due to reduced competition for target cells.

When the drug efficacy is time varying, the establishment probability as a function of dosing intervals is constrained on both ends (very frequent and infrequent dosing) by the probabilities associated with the time-averaged drug efficacy and the initial drug efficacy on account of the existence of the establishment time-frame (ETF) (**Figures S3C,E**). Depending upon whether the initial efficacy is lower (higher) than the time-average, the establishment probability decreases (increases) as the dosing interval increases. The maximum possible effect of dosing intervals is equal to the difference between the two constraining probabilities Δ_est_ = *P*_est_(*ϵ*(*t* = 0)) − *P*_est_(*ϵ*_avg_). The effect of increasing the dosing interval therefore amounts to moving along the *P*_est_ vs average drug efficacy curve. Positive Δ_est_ implies that long-acting drugs have a higher *P*_est_ and negative Δ_est_ implies the opposite. Given the nature of the curve (**Figure S3A**), parameters that increase Δ*ϵ* = |*ϵ*_avg_ − (*ϵ*(*t* = 0)| increase the effect of changing the dosing interval on *P*_est_, that is, lead to a higher |Δ_est_|. We’ve parameterized the slow absorbing drug model by the time-averaged drug efficacy *ϵ* _avg_ and the amplitude *A*,

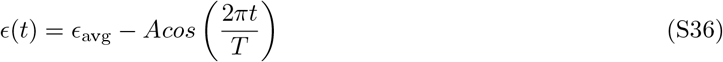

For this model, Δ*ϵ* = *A* and increasing the amplitude while keeping *ϵ*_avg_ fixed increases Δ_est_ (**Figure S3C**). Furthermore, due to the monotonicity of the curve, for such slow absorbing drug models the establishment probability will always be lower for longer-acting therapies compared to daily dosing as *ϵ*(*t* = 0) *< ϵ*_avg_.

In the fast absorbing drug model the efficacy is given by,

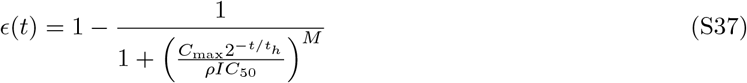

The dependence of treatment parameters on Δ_est_ is bit more complicated as any parameter combination that increases Δ*ϵ* corresponds to a larger |Δ_est_|. One way to do this is by varying the half-life *t*_*h*_ of the drug (**Figure S3E**) keeping *E*(*t* = 0) fixed. The effect is more pronounced for drugs with a shorter half-life. In contrast to the slow absorption model, the establishment probability will always be higher for long-acting therapies as compared to daily dosing when drug absorption is fast (*ϵ*(*t* = 0) *> ϵ*_avg_).

#### Mutants partially resistant to treatment

Treatment induces two opposing selection forces on a partially resistant strain - direct suppression of the strain by the drug and an indirect beneficial effect due to WT suppression that leads to more availability of target cells. Depending upon their relative intensity, a treatment with higher efficacy can increase or decrease the establishment probability for the resistant strain. To illustrate this, we plot the establishment probability as a function of constant drug efficacy for varying degrees of resistance in **Figure S3B**. Due to the non-monotonic nature, increasing the drug efficacy doesn’t always increase the establishment probability. Depending upon the degree of resistance, the suppression of the resistant strain by treatment can overpower the benefit conferred to it by a strong WT suppression and lead to a much lower establishment probability (the region after the point of inflection on the curves).

The establishment probability as a function of the dosing intervals is again constrained on both ends by the probabilities associated with the time-averaged drug efficacy and the initial drug efficacy due to the ETF (**Figures S3D,F**). For the slow absorbing drug model, the amplitude *A* still controls the difference between the two limiting cases and a smaller amplitude corresponds to a smaller |Δ_est_| (**Figure S3D**). The difference between the fully resistant case is that |Δ_est_| is not the maximum possible difference due to dosing intervals anymore. This is on account of the more complicated trends for intermediate dosing intervals caused by the two opposing selection forces. Slow absorbing (*ϵ*(*t* = 0) *< ϵ*_avg_) long-acting drugs increase the risk of resistance when the direct suppression of the resistant strain is the dominant selection force (**Figure S3D**), in contrast to when competition with the WT for available target cells dominates (**Figure S3C**).

Similarly, for the fast absorbing drug model, increasing Δ*ϵ* increases |Δ_est_| (**Figure S3F**). Shorter half-life of the drug leads to a larger effect of the drug dosing intervals on the establishment probability. In this case, long-acting drugs (*ϵ*(*t* = 0) *> ϵ*_avg_) reduce the risk of resistance when the direct suppression of the resistant strain is the dominant selection force (**Figure S3F**). The opposite is true when competition with the WT for available target cells dominates (**Figure S3E**).

#### Role played by treatment parameters on the rescue probability

As mentioned in the main text, the rescue probability is always dominated by the residual rate of mutant production from the WT and hence, has a similar dependence on dosing intervals for both mutants fully and partially resistant to treatment. This is reflected in the rescue probability vs constant drug efficacy curves for varying degrees of resistance (**Figure S5A**). Treatment with higher efficacy is always worse for rescue mutants. For time-varying drug levels the rescue probability is also mainly influenced by the drug kinetics during the ETF, and so is constrained on both ends by the probabilities due to the time-averaged drug efficacy and the initial drug efficacy (**Figures S5B,C**). Depending upon whether the initial efficacy is higher (lower) than the average leads to a lower (higher) probability of rescue. As a result, slow absorbing (*ϵ*(*t* = 0) *< ϵ*_avg_) long-acting drugs will always have a higher risk of resistance, and fast absorbing (*ϵ*(*t* = 0) *> ϵ*_avg_) long-acting drugs will have a lower risk of resistance as compared to daily dosing.

Similar to the case of pre-existence we define Δ_resc_ = *P*_resc_(*E*(*t* = 0)) − *P*_resc_(*ϵ*_avg_). Positive Δ_resc_ implies that long-acting drugs have a higher *P*_resc_ and negative implies the opposite. Given the nature of the curves, parameters that increase Δ*ϵ* lead to a larger Δ_resc_; for example, increasing the amplitude in the slow absorption model (**Figure S5B**) and reducing the half-life in the fast absorption model (**Figure S5C**).

#### Role played by the infection parameters on the establishment probability of pre-existing mutants

The establishment probability of a pre-existing resistant mutant depends upon the infection parameters that control the rate of target cell dynamics, infected cell dynamics, and the cost of resistance. We plot the establishment probability as a function of constant drug efficacy with varying target cell production rates *λ* for both partially and fully resistant mutants (**Figures S4A and S4B**). *R*_0_ of the WT is kept fixed (ratio *λ/d* is fixed, where *d* is the target cell death rate) in order to isolate the effects of just changing the timescale of the target cell dynamics. The establishment probability increases with faster target cell dynamics. The resistant strain can establish infection sooner due to faster target cell availability, lowering the chance of stochastic extinction. We also vary the infected cell death rate *a* (**Figures S4C and S4D**) (*R*_0_ of the WT is kept fixed by varying the infectivity parameter *b* proportionally to *a*) and the cost of resistance *s* (**Figures S4E and S4F**). Slower death rate and a lower cost of resistance reduces the chance of stochastic extinction and increases the establishment probability.

As mentioned in the previous section, the effect of changing the dosing interval for drugs with a time-varying drug efficacy amounts to moving along these constant efficacy curves due to the ETF. For example, in **Figure S4A** knowing the initial and the time-averaged drug efficacy allows us to calculate the maximum possible difference in the establishment probability |Δ_est_| on increasing the dosing interval, and the direction in which it would go (*P*_est_(*ϵ*_avg_) → *P*_est_(*ϵ*(*t* = 0)). For fully resistant mutants, changing these parameters doesn’t change the monotonic nature of the curves and hence, long-acting therapy will always be associated with a reduced risk of resistance for slow absorbing drugs, and an increased risk for fast absorbing drugs. These curves are not monotonic for mutants that are partially resistant to treatment on account of the competing selection forces. However, the turning points of the curves are largely unaffected on varying these parameters — they are influenced mainly by the degree of resistance — suggesting that the regime in which each of the selection forces dominates is unchanged. As a result, varying these parameters will not affect the direction of change in the establishment probability that is observed for a given drug on increasing the dosing interval. It does however affect the magnitude of the change |Δ_est_| as the slopes of these curves are affected. A steeper slope corresponds to a larger effect on *P*_est_ on changing the dosing intervals; for example, the effect increases for faster target cell dynamics, and slower infected cell death rates.

#### Role played by the infection parameters on the rescue probability

We also investigate the dependence of the rate of target cell dynamics, infected cell dynamics and the cost of resistance on the rescue probability in **Figure S6**. Again, we keep *R*_0_ of the WT strain fixed in order to isolate the effect of just varying the parameters. Faster target cell dynamics (**Figure S6A**), slower death rate of infected cells (**Figure S6B**) and lower mutation costs (**Figure S6C**) lead to higher rates of rescue.

As for pre-existence, the effect of changing the dosing interval for a time-varying drug efficacy is dependent on the local curvature of the rescue probability vs constant drug efficacy lines (for example, **Figure S6A**). Given that these curves are monotonic irrespective of the degree of resistance of the mutant strain, long-acting therapy will lead to an increased risk of resistance due to rescue mutants for the slow absorbing drug (*ϵ*(*t* = 0) *< ϵ*_avg_), and a reduced risk for the fast absorbing drug (*ϵ*(*t* = 0) *> ϵ*_avg_). The magnitude of the effect |Δ_resc_|, which is reflected in the curvature, is higher for faster target cell dynamics, slower rates of infected cell death, and lower costs of resistance. Note that these trends are true apart from the regime where the establishment of a resistant infection is guaranteed irrespective of the drug efficacy. For example, *P*_resc_ ∼1 for the darkest lines in Figures S6A and S6B unless the drug efficacy is very high (*ϵ* ∼1). In this regime *P*_resc_ will be unaffected by the dosing frequency.

#### Results under the delayed adherence model

Comparing with the results for the random adherence model (**Figure 4**), we find that for pre-existing and rescue mutants (**Figure S7** subplots A,B,D,E) the trends due to non-adherence are unchanged – long-acting drugs are more robust to the effects of non-adherence. This is on account of the resistance risks being mainly dependent upon the drug kinetics during the establishment time-frame (ETF). The number of doses taken during this time-frame decreases on increasing the dosing interval, and therefore the chance of missing a dose during the ETF is lower. As the ETF exists independent of the details of the adherence model and the first dose is always taken, the trends are unaffected.

For the case of a persistent infection (**Figure S7** subplots C,F) we find that the risks of resistance due to non-adherence are elevated overall in the delayed adherence model as compared to the random adherence model. This is especially true for dosing intervals *T* ∼ > 60, and this can be understood as follows. These trends are a result of the combined effect of the rate of mutant production from the WT and competition between the two strains for uninfected cells once the resistant mutants have been produced. The risk of resistance is highest in the regime where missing a dose leads to a burst of WT replication and mutant production, but where there is enough WT suppression overall that a resistant strain can establish infection. In the random adherence model for *T* ∼ > 60 the risks of resistance were low because this regime was reached infrequently as after missing a dose there was enough time for the WT infection to rebound before the next dose is taken, making it harder for the mutant to establish infection. On the other hand, this regime is reached more frequently in the delayed adherence model, as allowing missed doses to be taken on subsequent days leads to fewer days when the WT is under suboptimal therapy. Consequently this reduces the chance of full WT rebound and allows the mutants that are produced to outcompete it and establish infection.

#### Fixation under time-varying selection pressures

Our analysis essentially amounts to calculating the fixation probability of a mutant under time-varying selection pressures. This problem has been studied in the population genetics literature, but is complicated to analyze unless the timescales of environmental change and fixation are very different. When environmental changes occur much faster than the time from mutant appearance to fixation, the time-averaged fitness of the mutant determines the fixation probability [92]. At the other extreme, when environmental changes occur slowly as compared to the timescale of selection, the fixation probability is governed by the environmental conditions at the time of the mutant introduction [93]. We observe exactly the same effects for drug resistance, with the “establishment time frame” corresponding to the time-scale of fixation and the drug kinetics reflecting environmental change. The time-average drug efficacy can capture the fate of the resistant mutant when dosing is frequent, whereas the initial drug concentration determines the risk of resistance for very long acting drugs. As demonstrated by Cvijovic et al [30], in the intermediate regime where the two time-scales are comparable, the dynamics of the environmental fluctuations (drug kinetics in our case) strongly influence the fixation of the mutant and can’t be explained by any effective selection pressure alone.

